# MR-KG: A knowledge graph of Mendelian randomization evidence powered by large language models

**DOI:** 10.64898/2025.12.14.25342218

**Authors:** Yi Liu, Joshua Burton, Winfred Gatua, Gibran Hemani, Tom R Gaunt

## Abstract

**Background:** The exponential growth of Mendelian randomization (MR) literature has created challenges for systematically organising and synthesising evidence, with key information fragmented across heterogeneous publications. We present MR-KG, a knowledge graph resource using large language models (LLMs) to systematically extract and structure published MR evidence at scale.

**Methods:** We evaluated eight OpenAI and local LLMs for extracting structured information from MR study abstracts. Two reviewers independently assessed extraction quality across 100 randomly selected studies. We applied the pipeline to over 15,000 MR studies published between 2003 and 2025, implementing trait profile similarity matching to identify related research questions and evidence profile similarity matching to assess concordance across studies.

**Results:** The LLM extractors achieved high scores across all assessment dimensions, and the resulting MR-KG resource provides comprehensive coverage of the MR literature. Semantic similarity matching revealed that whilst researchers explore conceptually related domains, they typically examine distinct exposure-outcome pairs. Temporal analysis documented substantial increases in trait diversity per study and significant improvements in reporting completeness following STROBE-MR guidelines. Reproducibility analysis showed that whilst a majority of replicated trait pairs achieve high concordance, a non-trivial minority showed discordant results, with substantial domain-specific variation. In addition, semantic matching quality varied substantially across disease categories.

**Conclusions:** LLM-based extraction can address information overload in MR research by extracting structured, queryable knowledge from fragmented literature. MR-KG enables systematic evidence synthesis, reveals domain-specific reproducibility patterns, and provides a continuously updated resource for the research community. This approach is generalisable to other research domains facing similar challenges.

## 1 Introduction

Mendelian randomization (MR) uses genetic variants as instrumental variables to investigate causal relationships between modifiable exposures and health outcomes [1, 2], with substantial growth in published literature over the last two decades [3, 4, 5]. By exploiting Mendel’s laws of inheritance, MR minimises confounding from environmental factors and reverse causation [6, 7, 8], offering unconfounded causal estimates whilst circumventing ethical and logistical constraints of interventional studies [9]. This growth has been facilitated by increasing GWAS availability through repositories [10, 11], methodological advances [12, 13, 14, 15, 16], as well as accessible software packages [17, 18].

However, this growth creates *information overload*, particularly in systematic organisation and synthesis of evidence [3]. No comprehensive mechanism exists to aggregate findings across studies or systematically assess knowledge states for specific exposure-outcome relationships, and so researchers must manually search and synthesise findings across massive volumes of publications, a process that is time-consuming, error-prone, and increasingly infeasible. This challenge is particularly acute for MR because studies examine overlapping exposures and outcomes using different genetic instruments, analytical methods, and populations [15, 19]. Lack of standardised reporting complicates these issues, as key information is often buried in heterogeneous text [20, 21]. Therefore, this *fragmentation of evidence* brought by information overload impedes evidence triangulation [22, 23] to integrate findings from MR analysis with other evidence to establish robust causal relationships.

Large language models (LLMs) offer automated, large-scale extraction of structured information from scientific literature, which have demonstrated capabilities in biomedical knowledge extraction [24, 25, 26, 27, 28, 29]. We address these challenges by developing MR-KG (https://epigraphdb.org/mr-kg), a knowledge graph resource using LLM-based extraction to systematically structure MR evidence at scale. We evaluated eight state-of-the-art LLMs for extracting structured information from MR study abstracts and applied the extraction pipeline to over 15,000 MR studies published between 2003 and 2025. Our pipeline implements trait profile similarity matching using semantic embeddings to identify conceptually related research questions and evidence profile similarity matching to quantify concordance across studies examining matched exposure-outcome pairs. Through case studies examining temporal trends and reproducibility patterns across disease domains, we demonstrate how automated extraction can datamine fragmented literature into queryable knowledge.

Our work relates to systematic MR evidence curation and biomedical knowledge graphs. Systematic reviews and meta-analyses provide syntheses for specific research questions [30, 31, 32, 33, 34]. Our previous works MR-Base [17] and OpenGWAS [11] provide foundational infrastructure for conducting new MR analyses, and EpiGraphDB [35] curates pre-computed MR results across exposure-outcome pairs ([36, 37, 38]). Other previous works also provide pre-computed MR results including DMRdb [39] and MRdb [40]. MR-KG can complement (but does not replace) these synthesised and pre-computed evidence resources by extracting and structuring MR evidence from published studies as reported by their authors, therefore providing literature-wide coverage spanning all published exposure-outcome pairs reported in the literature. MR-KG can also be linked with biomedical knowledge graphs and resources [41, 35, 42, 11, 43] via Experimental Factor Ontology mapping [44].

We first describe the LLM-based extraction methodology and MR-KG construction (Section 3), then present extraction performance evaluation and demonstrate MR-KG capabilities via two case studies (Section 4), and discuss implications for evidence synthesis (Section 5). Technical terms are explained in Section S1.

## 2 Data and methods

### 2.1 Extraction of structural MR evidence using LLMs

Abstracts were extracted from PubMed using the R package https://github.com/explodecomputer/pubmedplot. The search term ‘Mendelian randomization [tiab] OR Mendelian randomisation [tiab]’ was used to obtain all studies with the term in title or abstract.

For each study abstract, we instructed the LLM extractor to perform extraction tasks on five categories (A.1 …A.4, B) of information organised into two groups (A and B) in JSON format (See Section S2 for detail specifications and an example prompts set):

- Group A: Meta-information about the study, including:

**–** The exposure (A.1) and outcome (A.2) traits, which the LLM extractor categorised into 20 categories including molecular, socioeconomic, environmental, and anthropometric traits.
**–** The analytical methods (A.3), encompassing commonly used MR methods as well as other approaches such as negative controls, colocalization, sensitivity analysis, and triangulation that are frequently employed in MR publications.
**–** The population(s) from which study samples were drawn (A.4).
- Group B: Reported results, where each entry comprises the following information:

**–** The exposure trait and outcome trait.
**–** Statistics including regression coefficients (beta), units, odds ratios, hazard ratios, 95% confidence intervals, standard errors, and P-values. Statistics that were not reported in the original text were recorded as null.
**–** Direction of the relationship, indicating whether the exposure “increases” or “decreases” the outcome.

We evaluated two LLM families (eight models total): 1) OpenAI models via API: o4-mini, GPT-4.1, GPT-4o, GPT-5-mini, and GPT-5; and 2) open-source models deployed locally using Hugging Face Transformers [45]: Llama 3, Llama 3.2, and DeepSeek R1. Local models, using variants of the open-source model architectures with appropriate parameter sizes, were deployed on UK AIRR HPC Isambard-AI [46] in H100 GPUs. Whilst LLMs with larger parameter sizes can be expected to perform better overall, we were also interested in examining performance across specific aspects of the extraction, as smaller models may demonstrate comparable performance in certain tasks and are computationally less expensive to run.

Two reviewers (J.B and W.G) scored extracted data from 100 randomly selected studies. The assessment examined five extraction categories (A.1 …A.4, B), each evaluated on three dimensions:

1. *Accuracy*: are extracted results accurate?
2. *Detail*: are results described in appropriate detail?
3. *Completeness*: is information missing?

Each dimension comprised three specific questions (Table 1), totalling 46 questions per model.

**Table 1:** LLM extraction assessment questionnaire structure. The questionnaire comprises three main assessment areas evaluating accuracy, detail, and completeness of extracted data. Each area includes both quantitative metrics and qualitative assessments scored by two independent reviewers across 100 randomly selected studies.

| Question ID | Response Type | Question |
| --- | --- | --- |
| *Accuracy (Q-X-a)* |  |  |
| Q-X-a-1 | Number | Total number of results reported by model |
| Q-X-a-2 | Number | Number of results reported incorrectly |
| Q-X-a-3 | Score (1–10) | Overall assessment of accuracy of results |
| *Detail (Q-X-b)* |  |  |
| Q-X-b-1 | Number | Number of results reported with sufficient detail |
| Q-X-b-2 | Score (1–10) | Overall assessment of appropriate detail |
| Q-X-b-3 | Free text | Description of issues |
| *Completeness (Q-X-c)* |  |  |
| Q-X-c-1 | Number | Number of results not reported by model |
| Q-X-c-2 | Score (1–10) | Overall assessment of missing information |
| Q-X-c-3 | Free text | Description of results not reported by model |

Note: The questionnaire was applied to evaluate extracted data from each of the 8 LLMs across 5 extraction groups (X): Q-1 (Exposure Traits), Q-2 (Outcome Traits), Q-3 (Analytical Methods), Q-4 (Populations), Q-5 (Reported Results). Quantitative questions (Q-X-a-1, Q-X-a-2, Q-X-b-1, Q-X-c-1) collect count data, qualitative scores (Q-X-a-3, Q-X-b-2, Q-X-c-2) use a 10-point scale where higher scores indicate better performance, and free text questions (Q-X-b-3, Q-X-c-3) capture detailed observations. In addition, there is an extra question on the overall performance of the model for the extraction tasks on the abstract of a study.

### 2.2 MR-KG resource construction

#### 2.2.1 Trait profile similarity

Two studies investigating similar exposures, outcomes, or both are likely to address related research questions, and therefore we can identify related studies based on the overall semantic alignment of their *trait profiles* to enable matching even when studies use different terminology (e.g. “body mass index” versus “BMI”) or related but distinct concepts. This approach is particularly valuable for studies investigating multiple traits, as trait profile-level matching captures broader research focus and identifies studies with similar multi-domain investigation patterns, whereas individual trait-to-trait matching would only identify studies sharing specific exposures or outcomes.

Here we formalise the trait profile similarity between two studies as the average of maximum cosine similarities between the embeddings of the trait profiles of the two studies. Given two trait profiles *A* = {*a*_1_*, a*_2_*, …, a_n_*} and *B* = {*b*_1_*, b*_2_*, …, b_m_*} with corresponding embedding vectors v*_ai_* and v*_bj_*, the semantic similarity is calculated as:

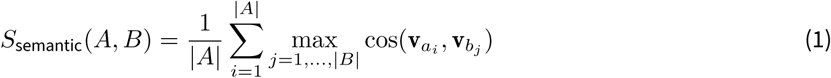

Where |*A*| denotes the cardinality (number of elements) in set *A* and 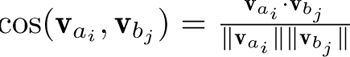 is the cosine similarity between embeddings. This metric ranges from 0 to 1, where 1 indicates perfect semantic alignment.

The semantic similarity, computed by averaging maximum cosine similarities between trait embeddings, captures conceptual overlap even when studies use different terminology. Complementing this, we also compute Jaccard similarity (size of intersection divided by size of union of trait index sets), which provides a conservative set-based measure with exact trait matches independent of embedding quality. Here, trait indices are unique integer identifiers assigned to each distinct trait label during preprocessing. For each query study, we compute both similarities against all other studies extracted using the same model to ensure extraction consistency. We rank candidate studies by their semantic similarity scores and retain the top 10 most similar studies for each query, whilst recording both metrics to enable analyses that consider embedding-based and set-based perspectives.

#### 2.2.2 Evidence profile similarity

In addition to trait profile similarity, which identifies studies investigating similar research questions, we calculate *evidence profile similarity* between studies, which examines the alignment of their observed evidence patterns. This distinction is important as studies with identical trait profiles may report concordant or discordant results depending on methodological factors, population characteristics, or true effect heterogeneity. Evidence profile similarity requires exactly or semantically matched exposure-outcome pairs between studies and assesses how their findings align in terms of effect directions, and when available in the abstracts, effect magnitudes and statistical significance. Unlike trait-level matching, which captures research focus overlap, evidence-level comparison evaluates finding reproducibility and can reveal systematic patterns or methodological factors affecting result concordance across studies.

Multiple metrics can quantify evidence profile similarity, including effect size correlation, significance agreement, and *direction concordance*, i.e. agreement in classified effect directions across matched exposure-outcome pairs. In MR-KG, we prioritise direction concordance as primary metric because structural data extracted from abstracts typically include effect directions with high reliability, whereas quantitative effect sizes, units and confidence intervals are less consistently reported. Given two studies *A* and *B* with *n* matched exposure-outcome pairs, where *d_A_*^(*i*)^*, d_B_*^(*i*)^ ∈ {−1, 0, +1} represent the classified directions (negative, null, positive), the direction concordance is calculated as:

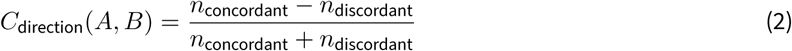

Where *n*_concordant_ is the number of pairs where both have same non-zero direction and *n*_discordant_ is the number of pairs where directions are opposite (one positive, one negative). Pairs involving null directions are excluded from calculation. This metric ranges from −1 (perfect discordance) to +1 (perfect concordance), with 0 indicating an equal number of concordant and discordant pairs.

Computing evidence profile similarity involves: 1) harmonisation of effect measures; 2) matching of exposure-outcome pairs between studies; and 3) calculation of similarity metrics. Studies are compared regardless of their overall trait profile similarity scores. For each pair of studies extracted using the same model, we first match exposure-outcome pairs by their trait labels to identify overlapping research findings. Matching proceeds through a hierarchical fallback scheme: first by exact trait index matches with identical normalised labels, and then by fuzzy matches via cosine similarity between semantic embeddings. We then harmonise effect sizes to a common beta (log) scale by transforming odds ratios and hazard ratios as *β* = log(OR) or *β* = log(HR) whilst keeping beta coefficients unchanged. For direction concordance specifically, we classify each harmonised effect as positive (*β >* 0),negative (*β <* 0), or null (*β* = 0 or missing), and compute concordance across all matched pairs. This categorical classification based on point estimate sign does not account for statistical uncertainty and in Section S3.1 we discuss important limitations of this approach. Beyond direction concordance, we also pre-compute other metrics when sufficient data are available (Section S3.1), including effect size correlation and statistical consistency, though the availability of these is substantially lower due to incomplete reporting in abstracts.

### 2.3 MR-KG compute infrastructure

As illustrated in Figure 1, the MR-KG resource comprises a set of DuckDB SQL databases, which curates the LLM extraction data and supported by other pre-computed artefacts (e.g. trait and evidence profiles as well as similarity mappings). For text embeddings of trait terms and EFO terms we used ScispaCy [47] (en_core_sci_lg-0.5.4). We also implemented auxiliary components including a web application and an API service for interactive and programmatic queries.

**Figure 1:**
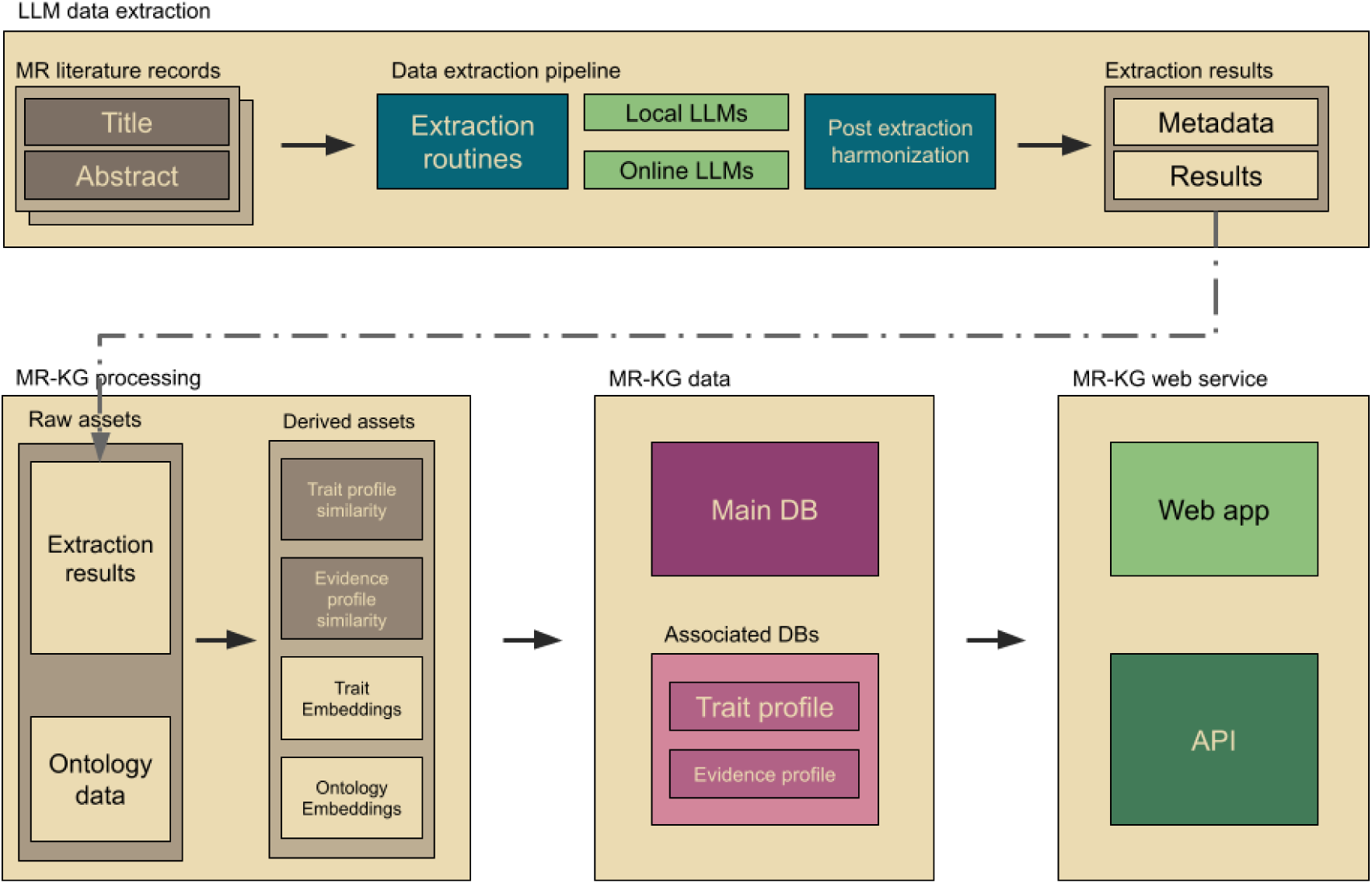
Architecture of the MR-KG system for automated extraction and knowledge graph construction. The system comprises two integrated components. Top panel: LLM data extraction pipeline processes MR literature records (titles and abstracts) through extraction routines using both local LLMs and online LLMs from OpenAI, followed by post-extraction harmonisation to produce standardised metadata and results. Bottom panel: MR-KG processing workflow transforms raw assets (extraction results and ontology data from the Experimental Factor Ontology) into derived assets including trait and ontology embeddings computed via ScispaCy, and precomputed trait profile similarity and evidence profile similarity metrics. These assets are curated into DuckDB SQL databases comprising a main database and associated databases for trait profiles and evidence profiles, which are then served through a web application and API service. See Section 3.1 for extraction methodology and Section 3.2 for detailed descriptions of similarity metrics.

## 3 Results

### 3.1 LLM extraction results

We visualised the assessment results on the extraction performance across LLMs in Figure 2, and report complete quantitative data in Tables S4-1–S4-3 as well as the variation in assessment scoring in Figures S4-1–S4-5. The results show GPT-5-mini and GPT-5 as top performers (overall means of 9.67 and 9.65, respectively), followed closely by GPT-4.1 and o4-mini (9.61 and 9.60), with Llama 3.2, GPT-4o, DeepSeek-R1, and Llama 3 performing comparably (9.30–9.57). The narrow range of overall scores (0.37 points) combined with larger standard deviations (1.14–1.86) indicates that study characteristics contributed more variation than model choice. Accuracy and detail scores were consistently high across all models (above 9.3), whereas completeness identified as the most challenging dimension, particularly for results extraction (Q-5: 8.2–9.1) and methodological information (Q-3: 8.2–9.4). Performance also varied by extraction task: trait extraction (Q-1, Q-2) showed consistently high scores across all models, whilst results extraction (Q-5) exhibited the greatest inter-model variability. Figures S4-1–S4-5 show high inter-reviewer agreement for accuracy assessments but greater variability in completeness scoring, which suggests that judgements about missing information are inherently more subjective. Comparison between model families showed that OpenAI models perform marginally better than local models, although Llama 3.2 achieved competitive performance on several extraction tasks.

**Figure 2:**
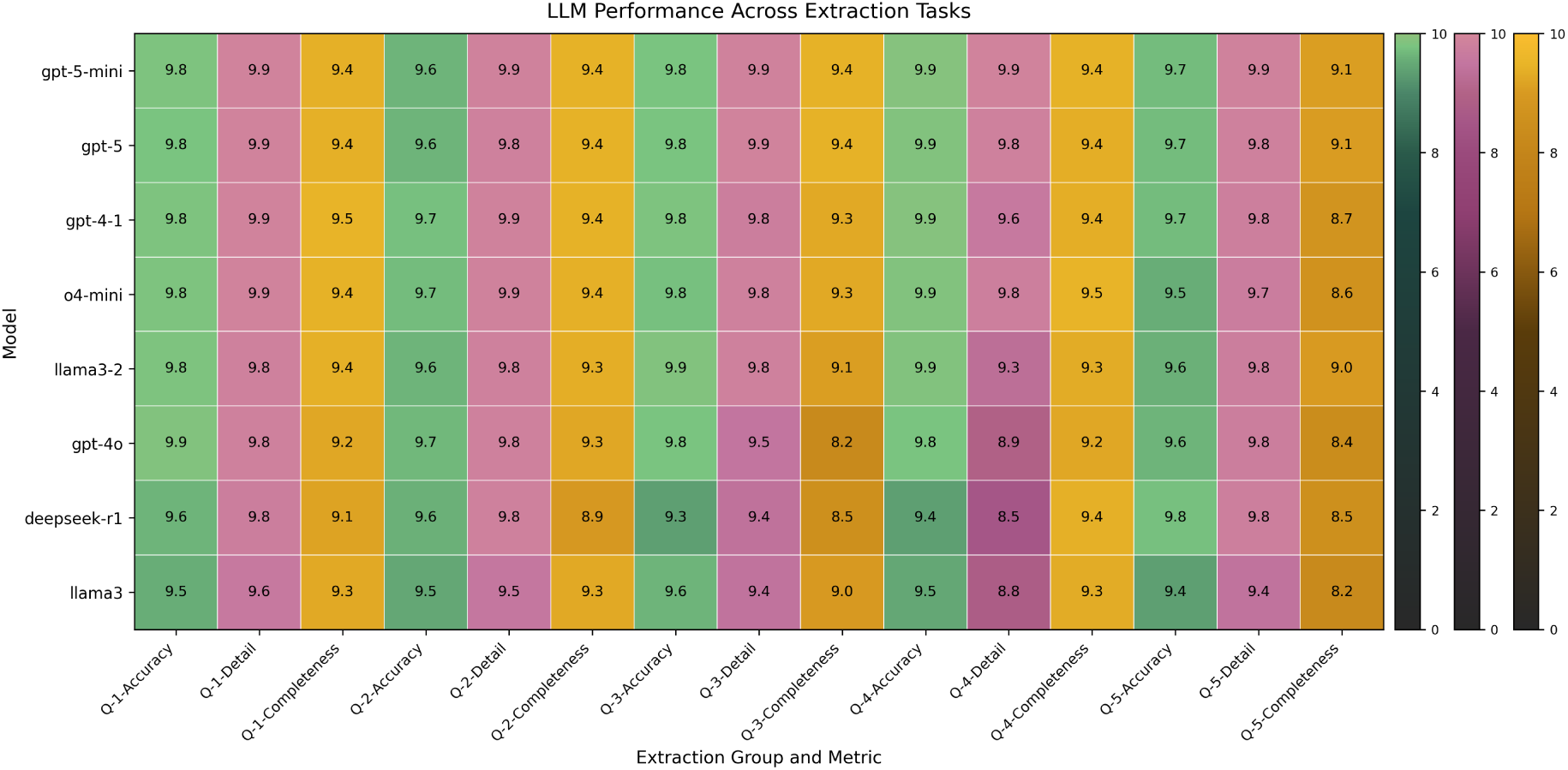
LLM extraction performance across task dimensions and extraction groups. Heatmap displaying mean assessment scores for 8 LLMs evaluated across 5 extraction groups and 3 assessment dimensions. Each cell value represents the mean of *overall*assessment scores (Q-X-a-3 for Accuracy, Q-X-b-2 for Detail, Q-X-c-2 for Completeness) from up to 200 assessments (100 studies × 2 independent reviewers), excluding missing scores indicating items not assessed. For each model and metric combination, scores were collected across 100 records and 2 reviewers (up to 200 assessments per cell), filtered to exclude zero scores indicating items not assessed, then aggregated using the arithmetic mean and rounded to 1 decimal place for display. Models ordered by overall performance. Extraction groups: Q-1 (Exposure Traits), Q-2 (Outcome Traits), Q-3 (Analytical Methods), Q-4 (Populations), Q-5 (Reported Results). Assessment dimensions: Accuracy (correctness of extracted information), Detail (appropriate level of specificity), Completeness (proportion of information captured).

Qualitative assessment of model outputs in the assessment sample showed substantial differences in schema compliance and data validity that were not captured by the scoring metrics. The human assessment also identified systematic patterns in model behaviour, including appropriate handling of bidirectional MR analyses, extraction of negative results, and management of missing information. OpenAI flagship models received explicit positive human assessments for robustness, with GPT-5 specifically noted for its capability in extracting evidence from text and handling negative result sentences, whilst local models exhibited interpretation issues including an inability to distinguish null effects from missing data and occasional hallucination of information not present in abstracts.

We additionally examined the schema compliance of the extracted results at scale across multiple LLMs. GPT-5 and GPT-4.1 have both demonstrated good compliance with the schema (Table S4-4), whereas other models have lower rates of compliance. Validation issue rates prevented automated processing for models with structural failures regardless of their content quality in subjective assessments. For downstream processing of the full MR-KG data, we used GPT-5 and GPT-4.1 to perform extraction tasks from the full MR literature corpus and present them as the default results in MR-KG. For other models, extracted data from GPT-5-mini and gpt-4o were omitted from data curation as we consider these models to be variants to their default models (GPT-5 and GPT-4.1) from the same architectures, but results from other models were included for comparison (Table S4-4).

### 3.2 MR-KG data resource

#### 3.2.1 Resource summary

As reported in Table 2, the MR-KG database aggregates extraction results from LLMs across 15,635 MR studies published between 2003 and 2025, yielding 253,650 individual MR results and a vocabulary of 75,121 unique trait labels. The default subset of GPT-5 extracted 89,280 MR results with an average of 5.72 results per paper as well as 42,575 unique trait labels, reflecting comprehensive phenotypic coverage and detailed causal relationship reporting in contemporary MR literature. Summary statistics for other LLM subsets can be found in Tables S5-1–S5-5.

**Table 2:** MR-KG Summary Statistics. Overall database characteristics, GPT-5 model extraction results, and similarity metrics.

| **Extraction results (all models)** |  |
| --- | --- |
| PMIDs processed | 15,635 |
| Total trait mentions | 283,537 |
| Total unique traits | 75,121 |
| Total model extraction records | 50,402 |
| Total extraction results | 253,650 |
| **Extraction results (GPT-5)** |  |
| Total extraction results | 89,280 |
| Total trait mentions | 106,346 |
| Total unique traits | 42,575 |
| Average results per paper | 5.72 |
| **Trait Profile Similarity (GPT-5)** |  |
| Total PMID-model combinations | 15,606 |
| Total pairwise comparisons | 156,060 |
| Semantic similarity (mean) | 0.724 |
| Semantic similarity (median) | 0.753 |
| Jaccard similarity (mean) | 0.096 |
| Jaccard similarity (median) | 0.056 |
| **Evidence Profile Similarity (GPT-5)** |  |
| Total PMID-model combinations | 1,498 |
| Total pairwise comparisons | 1,249 |
| Direction concordance (mean) | 0.413 |
| Direction concordance (median) | 1.000 |
| Data completeness (mean) | 0.623 |

*Note:* Statistics derived from LLM-based extraction across 15,635 MR studies published between 2003 and 2025. *Total unique traits* represents the deduplicated trait vocabulary (75,121), while *total trait mentions* (283,537) counts all occurrences including duplicates across studies. *Trait profile similarity* quantifies semantic and set-based overlap between studies based on extracted trait labels, and *evidence profile similarity* assesses concordance in reported effect directions and statistical evidence strength. *Semantic similarity* uses 200-dimensional ScispaCy embeddings with cosine distance, and *Jaccard similarity* measures set overlap. *Data completeness* represents the proportion of extracted results per paper containing valid quantitative effect estimates (beta coefficient, odds ratio, or hazard ratio) with classifiable effect direction.

For trait profile similarity metrics, the mean semantic similarity of 0.724 (median 0.753) indicates that studies tend to investigate conceptually related traits, however the mean Jaccard similarity of 0.096 (median 0.056) demonstrates limited exact trait overlap between studies. This suggests that whilst researchers explore related phenotypic domains, they typically investigate exposure-outcome combinations that are distinct from previous studies, which is consistent with the exploratory nature of MR research, as researchers seek novel causal relationships whilst building upon existing evidence through related but non-identical research questions. For computational efficiency, we retain only the top 10 most similar studies for each of the 15,606 query studies, yielding 156,060 pairwise comparisons. Evidence profile similarity metrics revealed moderate agreement in effect directions. Comparisons yielded mean direction concordance of 0.413 (median 1.000), suggesting clustering at perfect agreement with a subset showing substantial discordance (further analysis in Section 4.3.2).

#### 3.2.2 Individual study examples

To illustrate MR-KG at individual study level, we examine two cases: PMID 39836328 (Study 1; [48]) investigating ischemic heart disease and ovarian cancer subtypes, and PMID 40325806 (Study 2; [49]) investigating immune cell phenotypes and breast cancer risk. Data for both studies were retrieved from the web application (Figure 3) and are reported in Section S6 and summarised in Table 3.

**Figure 3:**
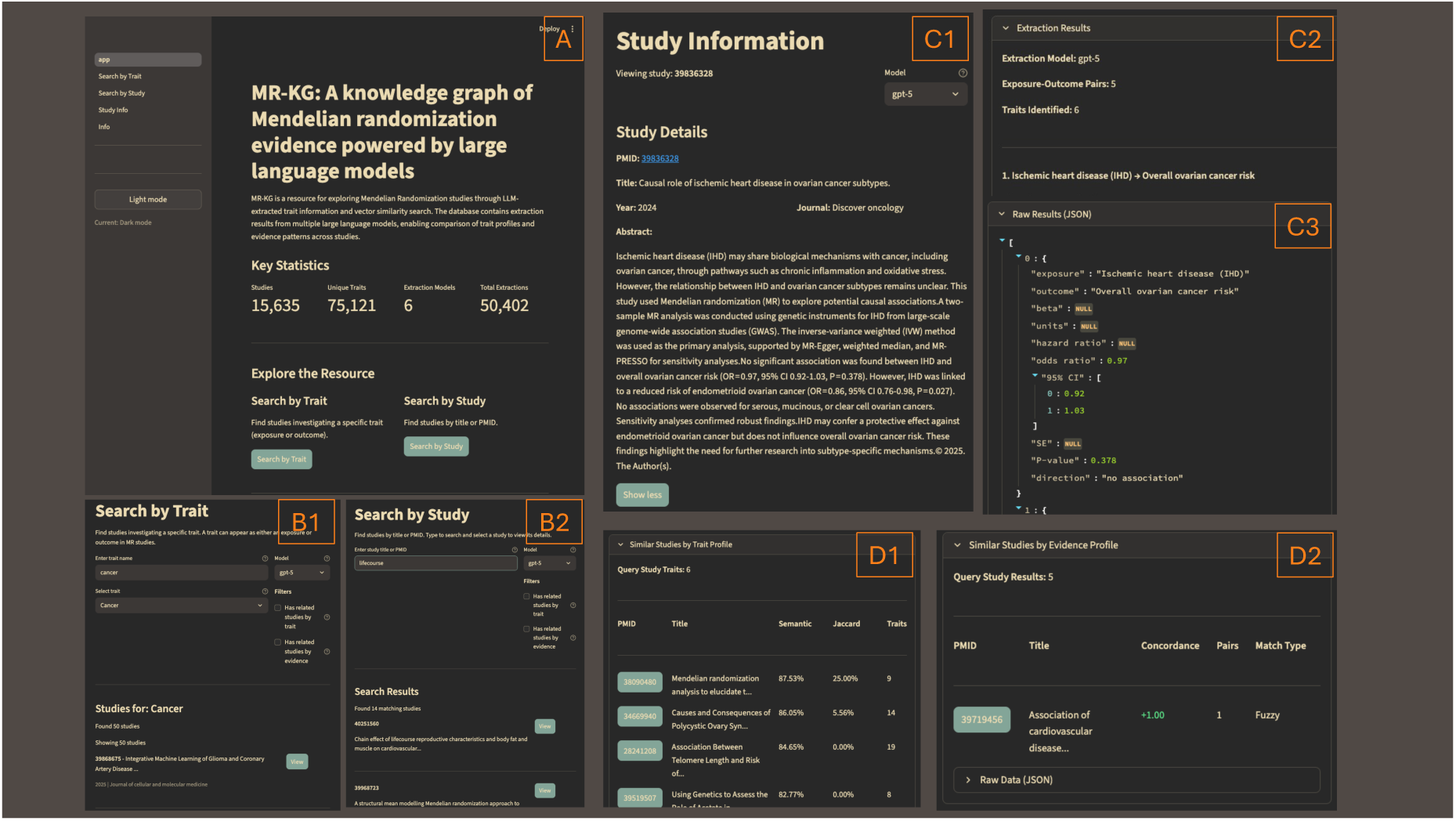
Functionalities of the MR-KG web application. **A**: Front page to the web application; **B**: Extracted data for studies can be accessed via searching for traits (B1) or search for the publication based on its title or PMID (B2); **C**: Curated data for studies, including its literature metadata (C1), extracted data as tables (C2) and raw JSON data (C3); **D**: Related studies to the query study, via trait profile similarity matching (D1) and evidence profile similarity matching (D2); The web application of MR-KG is available at https://epigraphdb.org/mr-kg. In addition, on the web application we also provide tutorials on programmatic queries to MR-KG via the API https://epigraphdb.org/mr-kg/api.

**Table 3:** Extracted data and related studies for two individual MR study examples. Comparison of LLM-extracted metadata, results, and similarity-based related studies for Study 1 (PMID 39836328; ischemic heart disease and ovarian cancer subtypes) and Study 2 (PMID 40325806; immune cells and breast cancer). Data extracted using GPT-5 model. Trait categories shown in square brackets.

| Field | Study 1 (PMID 39836328) | Study 2 (PMID 40325806) |
| --- | --- | --- |
| *Panel A: Extracted Metadata* |  |  |
| A.1 Exposures | Ischemic heart disease (IHD) [disease of the circulatory system] | Immune cell traits (731 phenotypes), CCR2 on monocytes, IgD- CD38dim AC cells, HLA-DR on dendritic cells, CD8dim NKT %T cells, CD8 on TD CD8bright cells, CD28+ CD45RA- CD8dim %CD8dim cells, plus 3 metabolite mediators [all molecular] |
| A.2 Outcomes | Ovarian cancer (overall risk), Endometrioid ovarian cancer, Serous ovarian cancer, Mucinous ovarian cancer, Clear cell ovarian cancer [all neoplasm] | Breast cancer [neoplasm] |
| A.3 Methods | Two-sample MR, sensitivity analysis, IVW, MR-Egger, weighted median, MR-PRESSO | Reverse MR, colocalization, sensitivity analysis, IVW, two-step MR, MR-Egger intercept test, Cochran's Q test |
| A.4 Population | Not specified in abstract | Immune cell traits (n=3,757), plasma metabolites (n=8,299), breast cancer (6,188 cases / 182,678 controls); ancestry not specified |
| *Panel B: Extracted Results* |  |  |
| Results extracted | 5 exposure-outcome pairs | 5 exposure-outcome pairs |
| Key findings | IHD → Overall ovarian cancer: OR=0.97 [0.92–1.03], P=0.378, no association; IHD → Endometrioid: OR=0.86 [0.76–0.98], P=0.027, decreases; IHD → Serous/Mucinous/Clear cell: no association (statistics not reported) | CCR2 on monocytes → BC: OR=0.969 [0.943–0.996], P=0.027, decreases; IgD- CD38dim AC → BC: OR=1.1 [1.023–1.183], P=0.009, increases; Metabolite mediators: directional effects only (statistics not reported) |
| Completeness | 2/5 pairs with full statistics (OR, CI, P-value); 3/5 with direction only | 2/5 pairs with full statistics; 3/5 with direction only (mediation results) |
| *Panel C: Related Studies* |  |  |
| **Trait Profile Similarity** |  |  |
| Top matches (semantic) | PMID 38090480 (sim=0.88): metabolites and ovarian cancer – shares Clear cell, Endometrioid, Ovarian cancer (overall); PMID 35492879 (sim=0.82): endometriosis and ovarian cancer histotypes – shares Clear cell, Endometrioid; PMID 28241208 (sim=0.85): telomere length and cancers – thematically similar (cancer outcomes) | PMID 39884473 (sim=0.70): immune cells and lung cancer/tuberculosis – shares immunophenotype methodology; PMID 39346920 (sim=0.70): immune cells and colorectal cancer – similar immune-cancer design; PMID 38826800 (sim=0.67): immune cells and breast cancer – shares Breast cancer outcome |
| **Evidence Profile Similarity** |  |  |
| Matched studies | 1 study: PMID 39719456 (cardiovascular disease and cancer) | 2 studies: PMID 38933268 (immune cells and pancreatic cancer); PMID 38935111 (immune cells and breast cancer) |
| Direction concordance | PMID 39719456: concordance=1.0 (1 fuzzy-matched pair); IHD → Endometrioid ovarian cancer (decreases) matched to Coronary heart disease → Breast cancer (decreases) – suggests generalised cardiovascular-cancer protective pattern rather than direct replication | PMID 38933268: concordance=0.0 (2 pairs); CCR2 on monocytes (decreases BC) vs CD11c+ monocytes (increases pancreatic cancer) discordant; IgD- CD38dim AC [B cell] (increases BC) vs HLA DR+ CD8br T cells [T cell] (increases pancreatic cancer) directionally concordant but different cell lineages. PMID 38935111: concordance=–1.0 (1 pair); IgD- CD38dim AC [B cell] (increases BC) vs CD3 on CD28+ CD4-CD8- T cells [T cell] (decreases BC) – reflects biological heterogeneity across immune cell lineages |

Note: BC = breast cancer; IHD = ischemic heart disease; IVW = inverse-variance weighted; MR = Mendelian randomization; OR = odds ratio; CI = confidence interval.

**Table 4:**
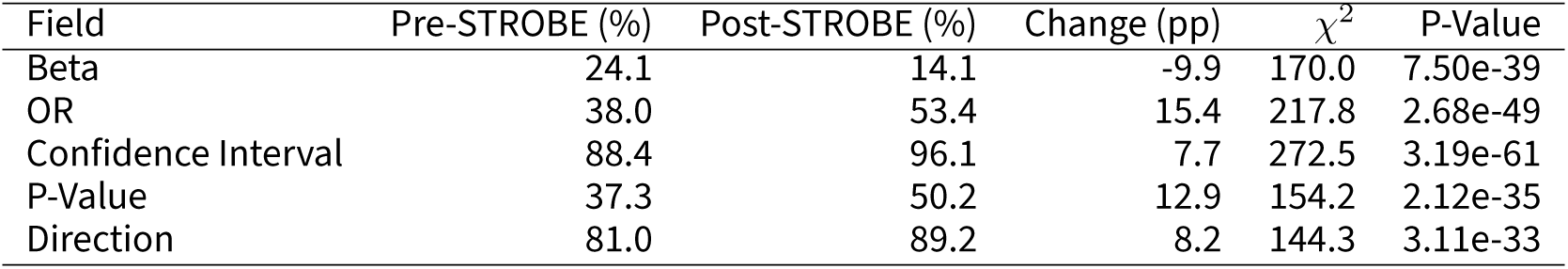
Impact of STROBE-MR guidelines (2021) on reporting completeness. Direct pre-post comparison centred on STROBE-MR publication year (2021). Pre-STROBE period includes studies published 2003–2020 (n=2,819); Post-STROBE period includes studies published 2021–2025 (n=12,780). Completeness percentages represent the proportion of studies reporting each field. Change is expressed in percentage points (pp). Statistical significance assessed using *χ*^2^ tests on 2×2 contingency tables comparing field presence/absence between periods.

| Field | Pre-STROBE (%) | Post-STROBE (%) | Change (pp) | $\chi^2$ | P-Value |
| --- | --- | --- | --- | --- | --- |
| Beta | 24.1 | 14.1 | -9.9 | 170.0 | 7.50e-39 |
| OR | 38.0 | 53.4 | 15.4 | 217.8 | 2.68e-49 |
| Confidence Interval | 88.4 | 96.1 | 7.7 | 272.5 | 3.19e-61 |
| P-Value | 37.3 | 50.2 | 12.9 | 154.2 | 2.12e-35 |
| Direction | 81.0 | 89.2 | 8.2 | 144.3 | 3.11e-33 |

For both studies, extractions accurately captured exposure and outcome traits, methods, and population information where available (Table 3). Results showed appropriate detail, with statistics recorded where reported and appropriately marked as null otherwise. Related studies retrieved via similar trait profiles were thematically appropriate, whilst those via similar evidence profiles suggest broader patterns of cardiovascular-cancer associations for Study 1 and highlight heterogeneity in immune cell-cancer findings for Study 2. For Study 1, the matched evidence suggests a generalised pattern of cardiovascular conditions having protective effects across cancer types rather than direct replication of the specific IHD-ovarian cancer relationship. For Study 2, the mixed and discordant evidence reflects genuine biological heterogeneity, as B cell and T cell phenotypes are distinct immune cell lineages that may exert opposing effects on cancer risk through different mechanisms.

### 3.3 Case studies

To demonstrate the analytical capabilities of MR-KG for systematic knowledge discovery and evidence synthesis, we present two case studies that illustrate how the resource can be utilised to address methodologically relevant questions in MR research. First, we examine temporal trends in MR research practices over two decades, assessing how trait diversity and reporting completeness have evolved. Second, we investigate the reproducibility and consistency of findings from independent MR studies investigating the same exposure-outcome relationships. These findings represent preliminary patterns identified through exploratory analysis, presenting opportunities for researchers using MR-KG for further investigation.

#### 3.3.1 Temporal trends in MR research

How has MR research evolved over the past two decades? Here we characterise the methodological development of MR through five successive eras defined by their representative landmark works, although some may fall outside the era boundaries (e.g. [16]). The *Early MR* era (2003-2014) established the conceptual and statistical foundations of MR as an instrumental variable approach [1, 2, 9]. The *MR-Egger* era (2015-2017) introduced MR-Egger regression [13] for detecting directional pleiotropy, spurring development of robust estimators including weighted median and mode-based methods [14, 19]. The *MR-PRESSO* era (2018-2019) systematised the identification and correction of horizontal pleiotropy [15, 50, 51]. The *Within Family MR* era (2020) introduced within-sibling designs to mitigate bias from population stratification, assortative mating, and dynastic effects [16, 52]. The *STROBE-MR* era (2021-2025) established reporting standards [53, 54], marking a shift from method invention to standardisation of analytical conduct and transparency. In order to assess how research practices evolved, we examined *trait diversity* (number of traits per study) and *reporting completeness* (proportion reporting key statistical elements in the abstract) across 15,606 studies (Table S7-1).

As shown in Figure 4, MR research has expanded substantially not only in volume but also scope: trait diversity increased substantially as MR studies now address broader phenotypic relationships. The mean number of traits per study increased from 4.11 (Early MR, n=454) to 7.14 (STROBE-MR, n=12,780) (Figure 4 B, Table S7-1). This progression was consistent across eras, indicating an evolution from single exposure-outcome investigations toward comprehensive phenome-wide approaches, likely enabled by increasing GWAS availability and analytical infrastructure.

**Figure 4:**
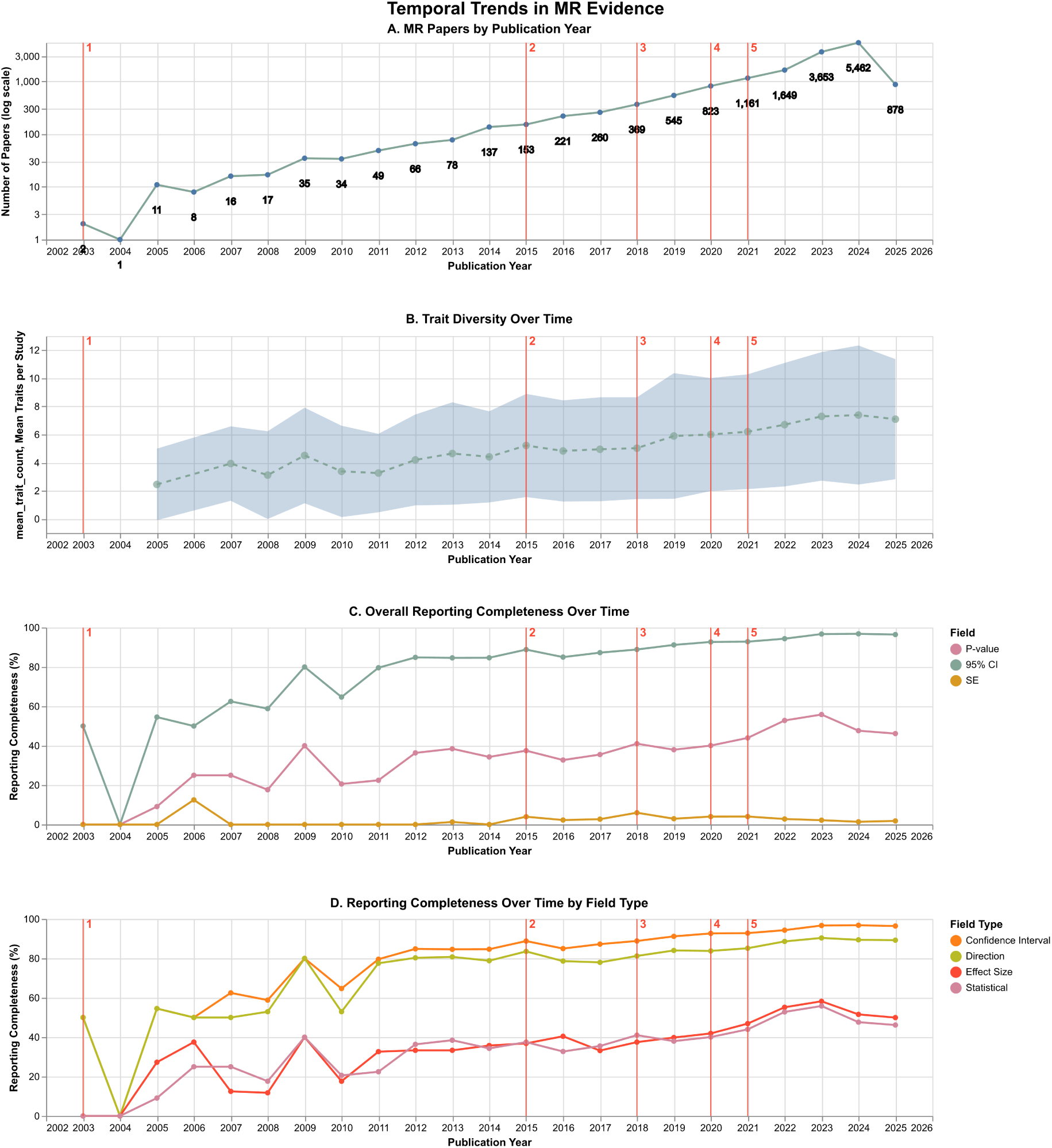
Temporal evolution of MR research (2003-04 – 2025-05). Multi-panel view showing longitudinal trends across five methodological eras (marked 1-5). **A**: Annual publication counts of MR research in our curated sample, plotted on log-scale for y-axis. **B**: Number of unique traits per study over time, showing steady increase in research scope. Trait diversity values for years 2003-2004 are not displayed due to insufficient observations. **C**: Overall reporting completeness percentage, demonstrating gradual improvement in methodological documentation. **D**: Field-type-specific completeness trends, revealing differential adoption patterns for quantitative effect estimates (OR, beta, CI) versus p-values and directionality. Era markers correspond to: (1) *Early MR* (2003-2014), (2) *MR-Egger* (2015-2017), (3) *MR-PRESSO* (2018-2019), (4) *Within-family MR* (2020), (5) *STROBE-MR* (2021-2025). Trend lines use locally weighted regression (LOESS) with shaded 95% confidence intervals.

Reporting practices in MR publications have improved substantially following the introduction of STROBE-MR guidelines, with all examined statistical elements showing significant increases (all p *<* 1e-30), suggesting the guidelines have had measurable impact on research communication standards. The proportion of studies reporting confidence intervals increased from 88.4 to 96.1 percent, achieving near-universal adoption. P-value reporting increased from 37.3 to 50.2 percent (though absolute levels remain lower), and effect direction reporting increased from 81.0 to 89.2 percent. Odds ratio reporting increased from 38.0 to 53.4 percent, implying greater emphasis on binary outcomes, whilst beta coefficient reporting decreased from 24.1 to 14.1 percent, possibly reflecting shifts toward standardised effect measures. Temporal trajectories (Figure 4 C-D) show upward trends across all field types with notable acceleration following 2021.

#### 3.3.2 Reproducibility and consistency of MR findings

When independent research teams investigate the same exposure-outcome relationships using MR analysis, do their findings agree? This question is fundamental to causal inference credibility, particularly as the MR literature spans multiple methodological eras and disease domains. We analysed 2,075 unique exposure-outcome trait pairs investigated by two or more independent studies to systematically evaluate reproducibility patterns.

We quantify *reproducibility* using direction concordance, which measures agreement in effect direction (positive, negative, or null) across pairwise comparisons within each trait pair. Analyses are stratified by trait *match type*, which reflects confidence in trait pair identity: exact matches (identical trait labels after normalisation) represent high-confidence pairs, whilst fuzzy matches (different labels matched via embedding similarity exceeding 0.70 threshold) represent pairs where semantic similarity suggests (but does not guarantee) investigation of the same biological relationship. We also stratify by *study count bands* (2-3, 4-6, 7-10, 11+ studies) and outcome category (cardiovascular, metabolic, psychiatric, cancer, autoimmune). Trait pairs are classified into *reproducibility tiers*: high (≥ 0.7), moderate (0.5-0.69), low (0.0-0.49), and discordant (less than 0.0).

Overall, 60.9 percent of trait pairs (n=1,264) achieve high reproducibility, whilst 8.4 percent (n=174) show moderate, 10.5 percent (n=218) show low, and 20.2 percent (n=419) demonstrate discordant results (Figure 5 A). Nearly 40 percent of relationships exhibit weaker than high-tier reproducibility, with one in five showing systematic disagreement. Reproducibility varies substantially across outcome categories (Figure 5 B). Psychiatric disorders exhibit the highest proportion of high-tier pairs (81.7 percent, n=94) with minimal discordance (9.6 percent). Cardiovascular and metabolic outcomes show intermediate patterns (64.5 percent and 64.6 percent high-tier, respectively). Cancer exhibits the lowest high-tier proportion (43.4 percent, n=169) with elevated discordance (24.4 percent), whilst autoimmune outcomes show moderate representation (59.4 percent high-tier) with notable discordance (24.5 percent). This 38 percentage point range between psychiatric and cancer domains suggests fundamental differences in instrument quality, phenotype precision, or biological complexity.

**Figure 5:**
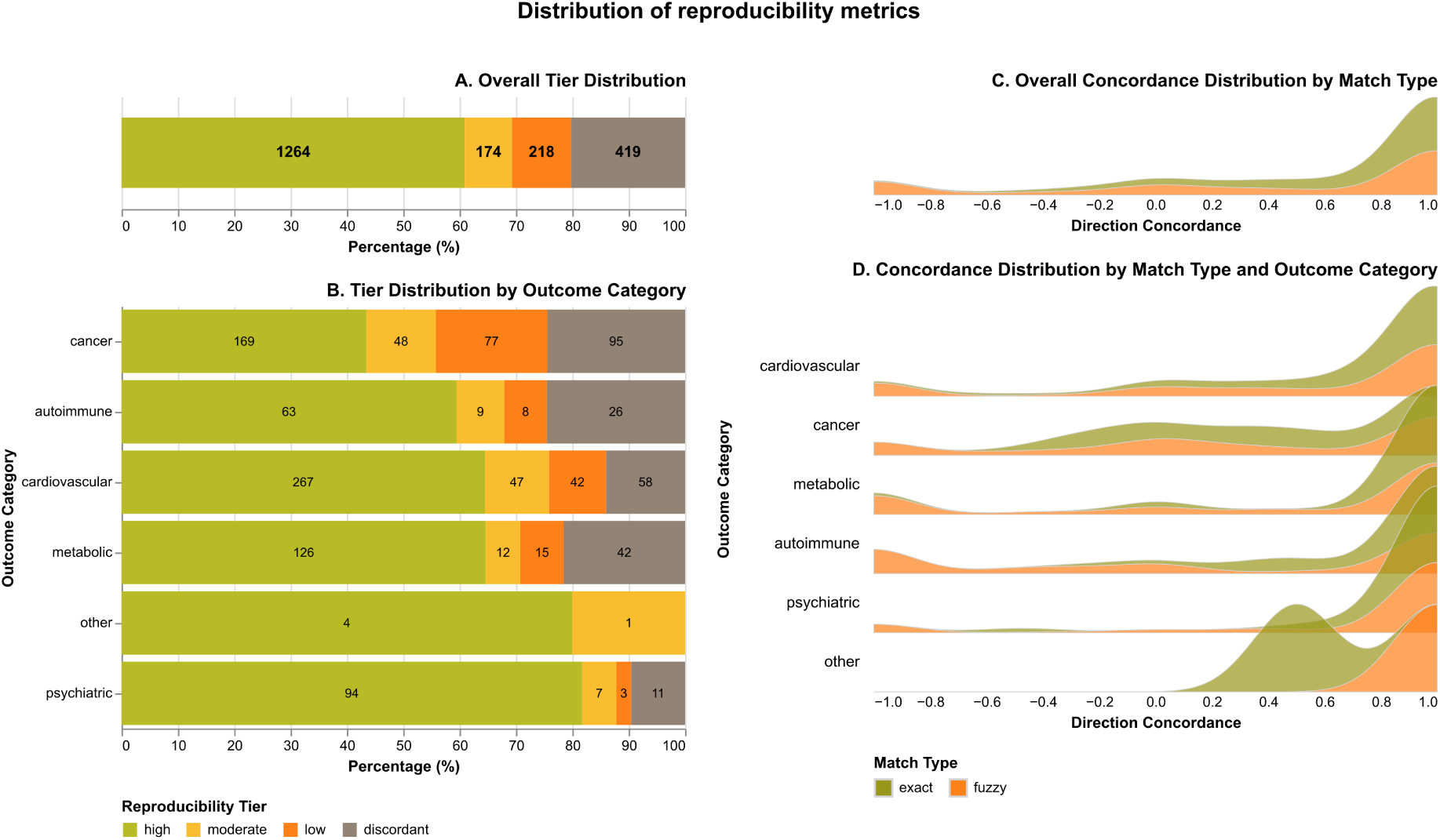
Distribution of reproducibility metrics across outcome categories and match types. Four-panel analysis showing reproducibility patterns organised in two complementary views: Left panel presents tier distributions for all trait pairs (**A**) and stratified by outcome category (**B**). Right panel presents concordance distributions using overlapping ridge plots stratified by match type for all trait pairs (**C**) and further stratified by outcome category (**D**). *Reproducibility tiers* defined as: High (direction concordance greater than or equal to 0.7), Moderate (0.5 to 0.69), Low (0.0 to 0.49), Discordant (less than 0.0). *Direction concordance* measures agreement in effect direction across studies: +1 if both report same direction, −1 if opposite directions, 0 if either is null. *Match types* represent confidence in trait pair identity: Exact (identical trait indices after normalisation, highest confidence), Fuzzy (pairs that failed exact matching but exceeded cosine similarity threshold of 0.70 on 200-dimensional trait embeddings, lower confidence). Of the 2,075 trait pairs, 1,224 were assigned to one of five outcome categories based on keyword matching; the remaining 851 pairs had outcome traits that did not match any predefined category keywords and were excluded from category-specific analyses.

Match type confidence substantially influences observed reproducibility (Figure 5 C-D, Table S7-2). Exact matches achieve mean concordance 0.739 (median 1.000, SD 0.441, n=261) whilst fuzzy matches show 0.414 (median 1.000, SD 0.760, n=963). The lower concordance and higher variability in fuzzy matches likely reflects both matching uncertainty (semantic similarity does not guarantee phenotypic equivalence) and genuine heterogeneity among related-but-distinct trait pairs. Autoimmune outcomes show large exact-fuzzy differences (0.670 units; exact: 0.843, fuzzy: 0.173), possibly reflecting conflation of terminologically similar but distinct conditions. Metabolic out-comes show substantial differences (0.479 units), cardiovascular shows moderate differences (0.255 units), whilst psychiatric disorders demonstrate robustness (0.178 units), possibly reflecting standardised diagnostic criteria. Cancer exhibits universally low concordance regardless of match type (exact: 0.462, fuzzy: 0.309), suggesting challenges beyond semantic matching including site-specific variation and genetic architecture differences. These patterns suggest phenotype matching strategies need tailoring to disease characteristics.

Study count exhibits a systematic relationship with concordance (Figure 6, Table S7-5): mean concordance declines from 0.50 (2–3 studies, n=1,492) to 0.44 (4–6 studies, n=437) to 0.33 (7–10 studies, n=105), with partial recovery to 0.44 (11+ studies, n=41). Regression analysis confirms a statistically significant negative association (beta = −0.024, p *<* 0.001, Table S7-3). A test for linear trend across ordered study count categories corroborates this finding (adjusted beta = −0.092, p *<* 0.001, Table S7-4), with non-parametric tests confirming the monotonic decline (Spear-man rho = −0.19, p *<* 0.001). Multiple mechanisms could produce this pattern: publication bias favouring concordant early replications, regression to the mean, and genuine heterogeneity accumulation as studies span diverse populations. The partial recovery in concordance at 11+ studies (from 0.33 to 0.44) suggests selective continuation of research, where trait pairs reaching high replication counts represent a filtered subset. Weak or inconsistent associations are likely abandoned after initial discordant results in the 7–10 study range, whilst associations sustaining 11+ replications may represent either truly robust effects or topics of sustained scientific interest despite mixed evidence. Concordance distributions shift from polarised (concentrated at extremes) in low study count bands to progressively dispersed with increasing replication. However, study count and match type together explain only 3.3 percent of the variance in concordance, indicating that reproducibility is primarily determined by trait pair-specific factors.

**Figure 6:**
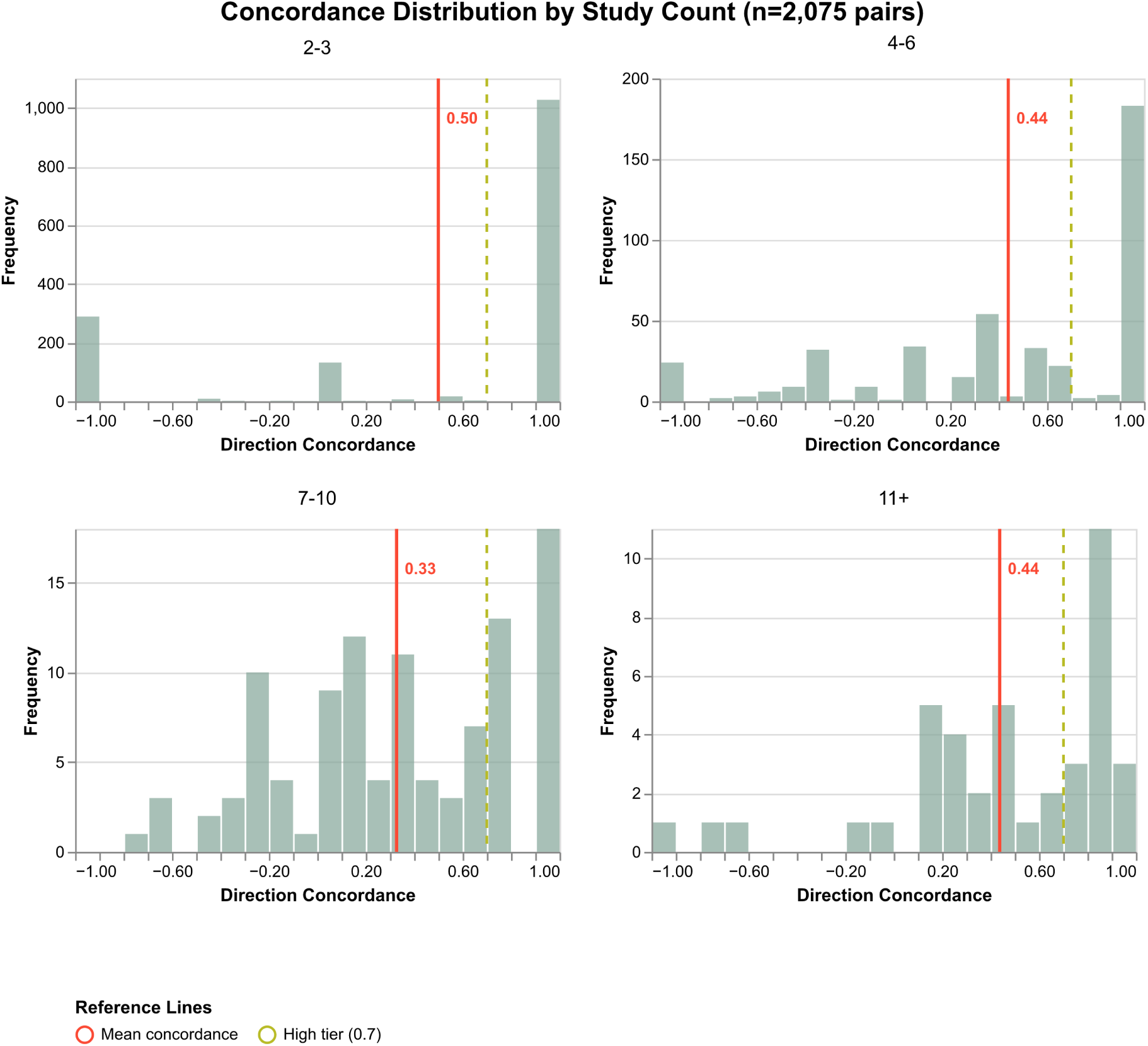
Impact of study count on cross-study reproducibility. Distribution of concordance scores across four study count bands (n=2,075 pairs), displayed as 2×2 faceted histograms. Each panel shows the frequency distribution of concordance values for trait pairs with different numbers of supporting studies: 2-3 studies (top left, n=1,492), 4-6 studies (top right, n=437), 7-10 studies (bottom left, n=105), and 11+ studies (bottom right, n=41). Direction concordance ranges from −1 (complete discordance) to +1 (perfect agreement), measuring directional agreement across pairwise study comparisons.

## 4 Discussion

We addressed the challenges of information overload and evidence fragmentation facing MR research by demonstrating that LLM-based extraction can systematically data-mine fragmented literature into structured, queryable knowledge. The MR-KG resource spans over 15,000 studies published between 2003 and 2025, encompassing more than 250,000 synthesised MR results. Two complementary similarity matching schemes enable knowledge discovery: trait profile similarity identifies studies investigating related research questions even when using different terminology, whereas evidence profile similarity quantifies consistency of findings across matched exposure-outcome pairs. MR-KG is publicly available at https://epigraphdb.org/mr-kg as a continuously updated resource for the research community.

Our case studies (Section 4.3) demonstrate the utility of structured evidence synthesis at scale powered by large language models. The temporal trends analysis documented the field’s evolution from focused single-relationship investigations toward comprehensive phenome-wide approaches, with trait diversity increasing by 73.8 percent and study volume growing from 454 to 12,780 across methodological eras. The temporal trajectory reveals a maturing field transitioning from methodological innovation (2003-2020) to standardisation and consolidation (2021-present), with STROBE-MR guidelines significantly improving statistical reporting, though the observational nature of this analysis cannot establish causality as concurrent factors may contribute to improved practices. The reproducibility analysis revealed that whilst a majority of repeatedly examined trait pairs achieve high concordance (60.9 percent), nearly 40 percent show weaker agreement including substantial discordance (20.2 percent), with considerable domain variation ranging from 81.7 percent high-tier in psychiatric disorders to 43.4 percent in cancer outcomes. These findings collectively demonstrate the fragmentation of MR evidence: whilst researchers explore conceptually related domains, they typically examine distinct exposure-outcome combinations, and comparisons across matched trait pairs show polarised concordance patterns. The individual study examples (Section 4.2.2) also illustrate this fragmentation: despite high semantic similarity in trait profiles, evidence profile matching identified only one to two comparable studies per query, reflecting the sparsity of directly replicable exposure-outcome pairs even within well-studied domains. In addition, the LLM extraction evaluation demonstrates that whilst flagship models achieved top performance, smaller ones showed competitive results on specific tasks: GPT-5-mini matched or exceeded GPT-5 performance, and open-source local models such as Llama 3.2 (11B parameters) achieved scores comparable to larger proprietary OpenAI models on trait extraction tasks despite being substantially smaller variants rather than flagship releases. Thus, there is considerable scope for domain-specific finetuned models with appropriately sized parameters to achieve competitive performance in evidence synthesis at reduced computational cost.

These findings directly address the core challenges of systematic MR evidence synthesis [3]. The substantial proportion of discordant findings as shown in our analysis empirically demonstrates why single MR studies should be interpreted cautiously and why evidence consistency across independent investigations is essential for robust causal conclusions. The trait profile similarity scheme enables identification of potentially redundant studies investigating well-established relationships, whilst simultaneously highlighting under-investigated exposure-outcome combinations where novel MR investigations would “genuinely advance knowledge” [3]. The domain-specific reproducibility patterns provide guidance for targeted synthesis strategies: psychiatric outcomes may benefit from flexible semantic matching given their robustness to match type, whilst autoimmune and metabolic domains may require stricter ontology-based harmonisation to avoid conflating distinct conditions, and cancer outcomes present particular challenges that extend beyond phenotype matching where biological knowledge and consideration (tumour site specificity, stage-dependent effects, etc.) would be required. The improved reporting completeness following STROBE-MR, particularly for confidence intervals and effect directions, enhances the feasibility of quantitative meta-analysis and evidence synthesis. By extracting key methodological details into a standardised schema, MR-KG provides the digital and methodological infrastructure necessary for systematic evidence synthesis, enabling researchers to move beyond individual study interpretation towards comprehensive assessment of MR evidence for wide-ranging causal questions.

As a proof-of-principle demonstration of automated MR evidence synthesis at scale, several limitations of the current MR-KG implementation represent opportunities for future research. Abstract-only extraction limits information completeness, as key details about genetic instruments, sensitivity analyses, and population characteristics are often reported exclusively in full text, and studies based on large numbers of exposures and/or outcomes (e.g. phenome-wide MR analyses) are unlikely to report all of these in the abstract. The use of direction concordance as the primary similarity metric was necessitated by inconsistent reporting of quantitative effect sizes. It is also important to note that directional discordance might not represent inconsistency between two results if these are close to the null with overlapping confidence intervals. Semantic similarity matching captures conceptual relatedness but cannot guarantee biological equivalence; our hierarchical matching scheme reflects this by assigning higher confidence to exact matches than to fuzzy matches, as evidenced by the domain-specific variation in direction concordance across match types. The resource also inherits publication bias from the underlying literature.

MR-KG opens multiple research directions. The most immediate one is systematic evidence triangulation [22], integrating MR-KG with RCT databases, observational cohorts, and mechanistic evidence to identify causal relationships supported across methodological approaches with non-overlapping sources of bias. Instrument quality assessment represents a second application, tracking consistency across studies using different genetic variants for the same exposure to reveal potentially problematic instruments affected by horizontal pleiotropy [13, 15]. The comprehensive trait coverage also enables phenome-wide investigations [55, 37] and systematic identification of research gaps. Finally, the approach is generalisable to other research domains facing information overload challenges, providing a template for automated evidence synthesis.

## Data Availability

Source code for LLM data extraction of MR studies is available at https://github.com/MRCIEU/llm-data-extraction. Source code for MR-KG infrastructure and analysis is available at https://github.com/MRCIEU/mr-kg. MR-KG data can be queried interactively via the web application https://epigraphdb.org/mr-kg and programmatically via the API https://epigraphdb.org/mr-kg/api.

## Acknowledgements

We used artificial intelligence (AI) tools as coding agents (further details reported in Section S8) to assist the implementation of code and documentation in research, as well as the scaffolding, initial outlining and final proof-editing of the manuscript. The current manuscript is fully written and revised by the lead author (Y.L.), and has been reviewed and edited by all authors. We have reviewed ISCB Policy for Acceptable Use of Large Language Models (https://www.iscb.org/iscb-policy-statements/iscb-policy-for-acceptable-use-of-large-language-models) and follow its guidelines on the acceptable uses.

## Funding

This work was funded by the UK Medical Research Council Integrative Epidemiology Unit (MC_UU_00032/1 and MC_UU_00032/3). This work was also supported by the National Institute for Health and Care Research (NIHR) Bristol Biomedical Research Centre; grant no: NIHR 203315. The views expressed are those of the authors and not necessarily those of the NIHR, the Department of Health and Social Care, or the Medical Research Council. The funders had no role in study design, data collection and analysis, decision to publish, or preparation of the manuscript. W.G is supported by a Wellcome Trust PhD studentship in Molecular, Genetic and Lifecourse Epidemiology (218495/Z/19/Z). This work was also supported by the University of Bristol Cancer Research Fund (UCRF) 2023 seedcorn funding “Assessing capability of large AI models in text mining of cancer studies”. The authors acknowledge the use of resources provided by the Isambard-AI National AI Research Resource (AIRR). Isambard-AI is operated by the University of Bristol and is funded by the UK Government’s Department for Science, Innovation and Technology (DSIT) via UK Research and Innovation; and the Science and Technology Facilities Council [ST/AIRR/I-A-I/1023].

## Conflicts of interest

T.R.G. receives funding from Biogen, Roche, GlaxoSmithkline and Novartis for unrelated research. Y.L receives funding from Roche for unrelated research.

