## supplementary-materials.pdf for "MR-KG: A knowledge graph of Mendelian randomization evidence powered by large language models"

### SUPPLEMENTARY MATERIALS FOR *MR-KG: A knowledge graph of Mendelian randomization evidence powered by large language models*

Yi Liu[1,*], Joshua Burton[1], Winfred Gatua[1], Gibran Hemani[1,2], and Tom R Gaunt[1,2,*]

[1] MRC Integrative Epidemiology Unit, Bristol Medical School, University of Bristol, Bristol, United Kingdom
[2] NIHR Bristol Biomedical Research Centre, University of Bristol, Bristol, United Kingdom

2025-12-14

#### S1 Glossary

Here we describe the various terms used in the manuscript and reference the main sections where these terms either first appear or are the topics for discussion.

- **Information overload** (Section 3.1) refers to the challenge facing the population health research community due to the exponential growth of MR literature, where systematic organisation and synthesis of the expanding evidence base becomes increasingly difficult. Researchers must manually search, read, and synthesise findings across massive volumes of publications, a process that is time-consuming, error-prone, and increasingly infeasible as the literature expands.
- **LLM assessment dimensions** (Section 3.1) are three sets of questions used to evaluate extraction performance:
  - *Accuracy*: whether extracted results are accurate.
  - *Detail*: whether extracted results are described in appropriate detail.
  - *Completeness*: whether there is any information not reported by the model.
- **Trait profiles** (Section 3.2.1) refer to the set of exposure and outcome traits investigated in a study. Two studies investigating similar exposures, outcomes, or both are likely to address related research questions, and MR-KG identifies related studies based on the overall semantic alignment of their trait profiles.
- **Evidence profiles** (Section 3.2.2) refer to the pattern of observed evidence in a study, comprising the set of exposure-outcome pairs and their associated quantitative findings. Evidence profile similarity assesses the alignment of these observed patterns between studies examining matched exposure-outcome pairs.
- **Trait profile similarity** (Section 3.2.1) quantifies the overlap in trait labels between studies to identify related research questions. The metric is calculated as the average of maximum cosine similarities between the embeddings of the trait profiles of two studies, capturing conceptual overlap even when studies use different terminology.
- **Evidence profile similarity** (Section 3.2.2) assesses concordance in quantitative MR findings between studies examining matched exposure-outcome pairs. This measure requires exactly or semantically matched exposure-outcome pairs and evaluates how findings align in terms of effect directions, and when available, effect magnitudes and statistical significance.
- **Direction concordance** (Section 3.2.2) is the primary metric used to quantify evidence profile similarity, measuring agreement in classified effect directions across matched exposure-outcome pairs. The metric ranges from $-1$ (perfect discordance) to $+1$ (perfect concordance), with 0 indicating an equal number of concordant and discordant pairs.
- **Match type** (Section 4.3.2) reflects confidence in trait pair identity when comparing studies. Exact matches use identical trait labels after normalisation (high-confidence pairs), whilst fuzzy matches use different labels matched via embedding similarity exceeding 0.70 threshold (pairs where semantic similarity suggests but does not guarantee investigation of the same biological relationship).
- **Reproducibility** (Section 4.3.2) is quantified using direction concordance to measure agreement in effect direction across pairwise comparisons within each trait pair investigated by two or more independent studies. Trait pairs are classified into *reproducibility tiers*: high ($\geq 0.7$), moderate (0.5–0.69), low (0.0–0.49), and discordant (less than 0.0).
- **Reproducibility tiers** (Section 4.3.2) are categories used to classify trait pairs based on their direction concordance scores: high ($\geq 0.7$), moderate (0.5–0.69), low (0.0–0.49), and discordant (less than 0.0). Overall, 60.9 percent of trait pairs achieve high reproducibility, whilst 20.2 percent demonstrate discordant results.
- **Trait diversity** (Section 4.3.1) refers to the number of distinct exposure and outcome traits investigated per study. MR research has shown substantial increases in trait diversity over time, with mean traits per study increasing from 4.11 (Early MR era) to 7.14 (STROBE-MR era), reflecting evolution toward comprehensive phenome-wide approaches.
- **Reporting completeness** (Section 4.3.1) measures the proportion of studies reporting key statistical elements in the abstract, including confidence intervals, P-values, effect directions, odds ratios, and beta coefficients. Reporting practices improved substantially following introduction of STROBE-MR guidelines, with all examined statistical elements showing significant increases.
- **Study count bands** (Section 4.3.2) stratify trait pairs by the number of independent studies investigating them (2–3, 4–6, 7–10, 11+ studies). Analysis revealed a systematic negative relationship between study

count and direction concordance, with mean concordance declining from 0.50 (2–3 studies) to 0.33 (7–10 studies), with partial recovery to 0.44 (11+ studies).

#### S2 Data extraction specifications

This section provides detailed specifications for the LLM-based extraction pipeline, including the prompting strategy, output schema, and computational infrastructure.

##### S2.1 Prompting strategy

We employed a multi-turn conversational prompting strategy to guide LLMs in extracting structured information from MR publication abstracts. The prompting pipeline consisted of four sequential components:

1. **System prompt**: Established the LLM's role as a data scientist responsible for extracting accurate information from research papers, with instructions to respond in JSON format.
2. **Abstract input**: Presented the study abstract to the LLM with contextual framing indicating it originated from an MR study.
3. **Example output with schema**: Provided a worked example demonstrating the expected output structure, accompanied by a formal JSON schema specification. This few-shot learning approach helped ensure consistent output formatting across different models.
4. **Task-specific extraction prompt**: Delivered detailed instructions for the extraction task, including:
   - Enumeration of all fields to extract
   - Predefined category lists for trait classification and method identification
   - Instructions for handling missing information
   - Output format requirements

For each abstract, two separate extraction calls were made: one for metadata extraction (exposures, outcomes, methods, and population) and one for results extraction (statistical findings). This separation allowed for specialised prompting tailored to each extraction task.

To ensure consistency in extracted data, we provided LLMs with predefined category lists for trait classification and method identification.

For exposure and outcome trait categorisation, we defined 20 categories based on established biomedical ontologies:

- Molecular
- Socioeconomic
- Environmental
- Behavioural
- Anthropometric
- Clinical measures
- Infectious disease
- Neoplasm
- Disease of the blood and blood-forming organs
- Metabolic disease
- Mental disorder
- Disease of the nervous system
- Disease of the eye and adnexa
- Disease of the ear and mastoid process
- Disease of the circulatory system
- Disease of the digestive system

- Disease of the skin and subcutaneous tissue
- Disease of the musculoskeletal system and connective tissue
- Disease of the genitourinary system
- Other (for traits not fitting the above categories)

For analytical methods, we defined 11 core categories commonly employed in MR studies:

- Two-sample Mendelian randomization
- Multivariable Mendelian randomization
- Colocalization
- Network Mendelian randomization
- Triangulation
- Reverse Mendelian randomization
- One-sample Mendelian randomization
- Negative controls
- Sensitivity analysis
- Non-linear Mendelian randomization
- Within-family Mendelian randomization

LLMs were instructed to match extracted methods to these predefined names where possible, and to specify “Other” along with the method name for approaches not in the list.

#### S2.2 Output schema

The extraction output was structured using JSON format with formal schema validation. We defined two complementary schemas: one for study metadata and one for reported results.

##### S2.2.1 Metadata schema

The metadata schema captured study-level information with the following structure:

```
{
  "metadata": {
    "exposures": [
      {"id": "<string>", "trait": "<string>", "category": "<string>"}
    ],
    "outcomes": [
      {"id": "<string>", "trait": "<string>", "category": "<string>"}
    ],
    "methods": ["<string>"],
    "population": ["<string>"],
    "metainformation": {
      "error": "<string>",
      "explanation": "<string>"
    }
  }
}
```

Each exposure and outcome entry included a unique identifier, the trait name as reported in the abstract, and the assigned category from the controlled vocabulary. The `metainformation` field allowed LLMs to document extraction challenges, such as missing population information or methods not matching the predefined list.

###### S2.2.2 Results schema

The results schema captured individual statistical findings:

```
{
  "results": [
    {
      "exposure": "<string>",
      "outcome": "<string>",
      "beta": <number|null>,
      "units": "<string|null>",
      "odds ratio": <number|null>,
      "hazard ratio": <number|null>,
      "95% CI": [<number|null>, <number|null>],
      "SE": <number|null>,
      "P-value": <number|null>,
      "Direction": "<increases|decreases>"
    }
  ],
  "resultsinformation": {
    "error": "<string>",
    "explanation": "<string>"
  }
}
```

LLMs were instructed to populate fields with `null` when values were not explicitly reported in the abstract. The `Direction` field captured the qualitative relationship between exposure and outcome, providing interpretable summary information even when numerical statistics were unavailable.

##### S2.3 Example prompts

Below we provide an example set of the extraction prompts.

##### S2.4 Computational infrastructure

###### Metadata extraction

```
[
    {
        "role": "system",
        "content": "You are a data scientist responsible for extracting accurate
information from research papers. You answer each question with a single JSON string.",
    },
    {
        "role": "user",
        "content": """
                This is an abstract from a Mendelian randomization study.
                "Alcohol consumption significantly impacts disease burden and has been
linked to various diseases in observational studies. However, comprehensive meta-analyses
using Mendelian randomization (MR) to examine drinking patterns are limited. We aimed
to evaluate the health risks of alcohol use by integrating findings from MR studies. A
thorough search was conducted for MR studies focused on alcohol exposure. We utilized
two sets of instrumental variables-alcohol consumption and problematic alcohol use-and
summary statistics from the FinnGen consortium R9 release to perform de novo MR analyses.
Our meta-analysis encompassed 64 published and 151 de novo MR analyses across 76 distinct
primary outcomes. Results show that a genetic predisposition to alcohol consumption,
independent of smoking, significantly correlates with a decreased risk of Parkinson's
disease, prostate hyperplasia, and rheumatoid arthritis. It was also associated with an
increased risk of chronic pancreatitis, colorectal cancer, and head and neck cancers.
Additionally, a genetic predisposition to problematic alcohol use is strongly associated
```

```
with increased risks of alcoholic liver disease, cirrhosis, both acute and chronic
pancreatitis, and pneumonia. Evidence from our MR study supports the notion that alcohol
consumption and problematic alcohol use are causally associated with a range of diseases,
predominantly by increasing the risk."
                """,
    },
    {
        "role": "assistant",
        "content": """This is an example output in JSON format:
    { "metadata": {
    "exposures": [
    {
        "id": "1",
        "trait": "Particulate matter 2.5",
        "category": "Environmental"
    },
    {
        "id": "2",
        "trait": "Type 2 diabetes",
        "category": "metabolic disease"
    },
    {
        "id": "3",
        "trait": "Body mass index",
        "category": "Anthropometric"
    }
    ],
    "outcomes": [
    {
        "id": "1",
        "trait": "Forced expiratory volume in 1 s",
        "category": "Clinical measure"
    },
    {
        "id": "2",
        "trait": "Forced vital capacity",
        "category": "Clinical measure"
    },
    {
        "id": "3",
        "trait": "Gastroesophageal reflux disease",
        "category": "disease of the digestive system"
    },
    {
        "id": "4",
        "trait": "Non-alcoholic fatty liver disease (NAFLD)",
        "category": "disease of the digestive system"
    }
    ],
    "methods": ["two-sample mendelian randomization", "multivariable mendelian
randomization", "colocalisation", "network mendelian randomization"],
    "population": ["European men", "Breast cancer patients", "African-Americans"],
    "metainformation": {
        "error": "No information on population is provided in abstract",
        "explanation": "Some methods do not match those listed in the prompt"
    }
    }
    }
    """,
```

```
    },
    {
        "role": "user",
        "content": """What are the exposures, outcomes in this abstract? If there are
multiple exposures or outcomes, provide them all. If there are no exposures or outcomes,
provide an empty list. Also categorize the exposures and outcomes into the following
groups using the exact category names provided:
- molecular
- socioeconomic
- environmental
- behavioural
- anthropometric
- clinical measures
- infectious disease
- neoplasm
- disease of the blood and blood-forming organs
- metabolic disease
- mental disorder
- disease of the nervous system
- disease of the eye and adnexa
- disease of the ear and mastoid process
- disease of the circulatory system
- disease of the digestive system
- disease of the skin and subcutaneous tissue
- disease of the musculoskeletal system and connective tissue
- disease of the genitourinary system
If an exposure or outcome does not fit into any of these groups, specify "Other".

List the analytical methods used in the abstract. Match the methods to the following list
of exact method names. If a method is used that is not in the list, specify "Other" and
also provide the name of the method. The list of methods is as follows:
- two-sample mendelian randomization
- multivariable mendelian randomization
- colocalization
- network mendelian randomization
- triangulation
- reverse mendelian randomization
- one-sample mendelian randomization
- negative controls
- sensitivity analysis
- non-linear mendelian randomization
- within-family mendelian randomization

Provide a description of the population(s) on which the study described in the abstract
was based.

Provide your answer in strict pretty JSON format using exactly the format as the example
output and without markdown code blocks. Any error messages and explanations must be
included in the JSON output with the key "metainformation".
""",
    },
]
```

#### Results extraction

```
[
    {
        "role": "system",
```

```
        "content": "You are a data scientist responsible for extracting accurate
information from research papers. You answer each question with a single JSON string.",
    },
    {
        "role": "user",
        "content": """
                This is an abstract from a Mendelian randomization study.
                "Alcohol consumption significantly impacts disease burden and has been
linked to various diseases in observational studies. However, comprehensive meta-analyses
using Mendelian randomization (MR) to examine drinking patterns are limited. We aimed
to evaluate the health risks of alcohol use by integrating findings from MR studies. A
thorough search was conducted for MR studies focused on alcohol exposure. We utilized
two sets of instrumental variables-alcohol consumption and problematic alcohol use-and
summary statistics from the FinnGen consortium R9 release to perform de novo MR analyses.
Our meta-analysis encompassed 64 published and 151 de novo MR analyses across 76 distinct
primary outcomes. Results show that a genetic predisposition to alcohol consumption,
independent of smoking, significantly correlates with a decreased risk of Parkinson's
disease, prostate hyperplasia, and rheumatoid arthritis. It was also associated with an
increased risk of chronic pancreatitis, colorectal cancer, and head and neck cancers.
Additionally, a genetic predisposition to problematic alcohol use is strongly associated
with increased risks of alcoholic liver disease, cirrhosis, both acute and chronic
pancreatitis, and pneumonia. Evidence from our MR study supports the notion that alcohol
consumption and problematic alcohol use are causally associated with a range of diseases,
predominantly by increasing the risk."
                """,
    },
    {
        "role": "assistant",
        "content": """This is an example output in JSON format:
    {
    "results": [
        {
            "exposure": "Particulate matter 2.5"},
            "outcome": "Forced expiratory volume in 1 s"
            "beta": 0.154,
            "units": "mmHg",
            "hazard ratio": null,
            "odds ratio": null,
            "95% CI": [0.101,0.215],
            "SE": 0.102,
            "P-value": 0.0015,
            "Direction": "increases"
        },
        {
            "exposure": "Body mass index"},
            "outcome": "Gastroesophageal reflux disease"
            "beta": null,
            "units": null,
            "hazard ratio": null,
            "odds ratio": 1.114,
            "95% CI": [1.021,1.314],
            "SE": null,
            "P-value": 0.0157,
            "Direction": "increases"
        },
        {
            "exposure": "Body mass index"},
            "outcome": "Non-alcoholic fatty liver disease (NAFLD)"
            "beta": null,
```

```
            "units": null,
            "hazard ratio": null,
            "odds ratio": null,
            "95% CI": [null,null],
            "SE": null,
            "P-value": null,
            "Direction": "increases"
        }
    ]
    "resultsinformation": {
        "error": "No results provided in abstract",
        "explanation": "P-values were string, not numeric values"
    }
    }
    """,
    },
    {
        "role": "user",
        "content": """
List all of the results in the abstract, with each entry comprising: exposure, outcome,
beta, units, odds ratio, hazard ratio, 95% confidence interval, standard error, and
P-value. If any of these fields is missing, substitute them with "null". Add a field
called "direction" which describes whether the exposure "increases" or "decreases" the
outcome.
Provide your answer in strict pretty JSON format using exactly the format as the example
output and without markdown code blocks. You must only include values explicitly written
in the abstract. Any error messages and explanations must be included in the JSON output
with the key "resultsinformation".

""",
    },
]
```

**OpenAI models** OpenAI models (o4-mini, GPT-4.1, GPT-4o, GPT-5-mini, and GPT-5) were accessed via the OpenAI Responses API. For reasoning-enhanced models (o4-mini), we configured the reasoning effort parameter to "medium" to balance extraction accuracy with computational cost. API calls were executed in batch mode with appropriate rate limiting to comply with service quotas.

**Local models** Local models (Llama 3, Llama 3.2, and DeepSeek R1) were deployed on the UK AI Research Resource (AIRR) High Performance Computer Isambard-AI at the Bristol Centre for Supercomputing (BriCS). Model inference was performed using the Hugging Face Transformers library with GPU acceleration. Extraction jobs were parallelized using SLURM array tasks, with each task processing a subset of abstracts to enable efficient scaling across the full corpus.

### S3 MR-KG specifications

#### S3.1 Evidence profile similarity metrics

The main text describes direction concordance as our primary metric for evidence profile similarity due to its universal availability. Here we provide specifications for supplementary metrics that quantify additional dimensions of evidence alignment when sufficient data are available.

**Effect size similarity.** Effect size similarity quantifies agreement in effect magnitude across matched exposure-outcome pairs using Pearson correlation of harmonised effect sizes. Given two studies $A$ and $B$ with $n$ matched exposure-outcome pairs, where $\beta_A^{(i)}$ and $\beta_B^{(i)}$ are the harmonised effect sizes for pair $i$, the effect size similarity is:

$$r_{\text{effect}}(A, B) = \frac{\sum_{i=1}^{n}(\beta_A^{(i)} - \bar{\beta}_A)(\beta_B^{(i)} - \bar{\beta}_B)}{\sqrt{\sum_{i=1}^{n}(\beta_A^{(i)} - \bar{\beta}_A)^2}\sqrt{\sum_{i=1}^{n}(\beta_B^{(i)} - \bar{\beta}_B)^2}} \tag{1}$$

where $\bar{\beta}_A$ and $\bar{\beta}_B$ are the mean harmonised effect sizes across matched pairs. This metric ranges from $-1$ (effect sizes systematically opposite) to $+1$ (effect sizes align perfectly), with $0$ indicating no correlation. Computation requires a minimum of 3 matched pairs with at least 2 unique effect size values in each study.

Abstracts frequently omit quantitative effect sizes, particularly for negative or null results, limiting the applicability of this metric. When available, this metric provides complementary information to direction concordance by capturing the magnitude of agreement beyond categorical direction.

**Statistical consistency.** Statistical consistency measures agreement in statistical significance classifications using Cohen's kappa coefficient. For two studies with $n$ matched pairs, where each pair is classified as significant ($p < 0.05$) or non-significant ($p \geq 0.05$), Cohen's kappa is:

$$\kappa = \frac{p_o - p_e}{1 - p_e} \tag{2}$$

where $p_o$ is the observed agreement proportion and $p_e$ is the expected agreement by chance. The expected agreement is calculated as:

$$p_e = p_{\text{sig},A} \cdot p_{\text{sig},B} + p_{\text{nonsig},A} \cdot p_{\text{nonsig},B} \tag{3}$$

where $p_{\text{sig},A}$ and $p_{\text{nonsig},A}$ represent the proportions of significant and non-significant pairs in study $A$ (and similarly for study $B$). This metric ranges from $-1$ (systematic disagreement, worse than chance) through $0$ (agreement equivalent to chance) to $+1$ (perfect consistency). Standard interpretation follows the Landis and Koch scale: $< 0.20$ slight, $0.21$–$0.40$ fair, $0.41$–$0.60$ moderate, $0.61$–$0.80$ substantial, and $> 0.80$ almost perfect agreement.

Statistical consistency has low availability from two compounding factors: first, p-values are inconsistently reported in abstracts; second, the metric requires a minimum of 3 matched pairs, but 82% of study pair comparisons share only 1 matched trait pair. Consequently, statistical consistency serves as an exploratory metric applicable only in rare cases where studies share multiple overlapping exposure-outcome pairs with complete significance data.

**Precision concordance.** Precision concordance measures similarity in effect estimate precision using Spearman rank correlation of confidence interval widths. For two studies with $n$ matched pairs, where $w_A^{(i)} = \text{CI}_{\text{upper}}^{(i)} - \text{CI}_{\text{lower}}^{(i)}$ represents the confidence interval width for pair $i$, the precision concordance is:

$$\rho = 1 - \frac{6\sum_{i=1}^{n} d_i^2}{n(n^2 - 1)} \tag{4}$$

where $d_i$ is the difference in ranks between $\log(w_A^{(i)})$ and $\log(w_B^{(i)})$. Log transformation is applied to handle the typically skewed distribution of confidence interval widths. Spearman rather than Pearson correlation is used to capture monotonic relationships without assuming linearity. This metric ranges from $-1$ (inverse precision patterns) to $+1$ (precision patterns align).

When available, this metric indicates whether studies achieve similar levels of precision across their shared exposure-outcome investigations, which may reflect comparable sample sizes, genetic instrument strength, or methodological rigour.

**Limitations of direction concordance classification.** Our direction concordance metric classifies effect directions categorically based on whether point estimates fall above or below zero. This approach has important methodological limitations that warrant consideration when interpreting concordance patterns.

First, the classification does not account for statistical uncertainty in effect estimates. Two studies reporting effect estimates on opposite sides of zero but with overlapping confidence intervals would be classified as discordant, even though they provide no statistical evidence of inconsistency. Such pairs represent compatible findings within sampling error rather than true discordance. Conversely, two estimates in the same direction but with non-overlapping confidence intervals may represent greater true discordance than opposite-signed estimates with substantial overlap. The categorical classification based on point estimate sign alone cannot distinguish these scenarios.

Second, the classification treats the null hypothesis (beta equals zero) as a point rather than a region. In practice, effect estimates very close to zero with wide confidence intervals may be classified as positive or negative based on minor sampling variation, when they more appropriately represent null or negligible effects. This issue is partially mitigated in our implementation by classifying effects as null when confidence intervals are available and span zero, but this information is inconsistently reported in abstracts.

Third, even when confidence intervals are available, abstract-level extraction introduces additional uncertainty. Confidence intervals may be incompletely reported, rounded, or presented in formats requiring transformation, each introducing potential misclassification. Combined with the categorical nature of direction classification, these extraction challenges mean our concordance metric provides a conservative lower bound on true statistical consistency rather than a definitive measure.

These limitations arise fundamentally from the constraints of abstract-level data extraction, where confidence intervals, standard errors, and p-values are inconsistently reported. Future work incorporating full-text extraction could enable more sophisticated consistency metrics that properly account for statistical uncertainty. Such approaches might include heterogeneity statistics commonly used in meta-analysis (e.g., Cochran's Q-test, I-squared), prediction interval overlap measures, or Bayesian methods for quantifying evidence consistency. Nevertheless, direction concordance remains valuable as a practical metric that leverages the most reliably extracted information from abstracts whilst enabling large-scale evidence pattern analysis across the MR literature.

#### S4 LLM performance on data extraction tasks

Table S4-1: **Comprehensive LLM extraction performance across task dimensions (overall, Q-1).** Mean assessment scores and standard deviations for 8 models evaluated across 5 extraction groups and 3 dimensions. Models ordered by overall mean performance. Overall mean and standard deviation calculated across all 15 group-dimension combinations per model. Extraction groups: Q-1 (Exposure Traits), Q-2 (Outcome Traits), Q-3 (Analytical Methods), Q-4 (Populations), Q-5 (Reported Results). Assessment dimensions: Accuracy (correctness), Detail (specificity), Completeness (coverage). Scores derived from 2 independent reviewers assessing 100 studies per model, with up to 200 assessments per cell. Zero scores (indicating items not assessed) excluded before calculating means.

| model | family | overall_mean | overall_sd | Q-1-Accuracy | Q-1-Completeness | Q-1-Detail |
|---|---|---|---|---|---|---|
| gpt-5-mini | OpenAI | 9.67 | 1.14 | 9.82 | 9.41 | 9.89 |
| gpt-5 | OpenAI | 9.65 | 1.18 | 9.79 | 9.41 | 9.89 |
| gpt-4-1 | OpenAI | 9.61 | 1.23 | 9.83 | 9.47 | 9.90 |
| o4-mini | OpenAI | 9.60 | 1.27 | 9.84 | 9.44 | 9.91 |
| llama3-2 | Local | 9.57 | 1.24 | 9.84 | 9.39 | 9.81 |
| gpt-4o | OpenAI | 9.40 | 1.58 | 9.85 | 9.25 | 9.84 |
| deepseek-r1 | Local | 9.30 | 1.78 | 9.63 | 9.15 | 9.75 |
| llama3 | Local | 9.30 | 1.86 | 9.54 | 9.28 | 9.59 |

Table S4-2: **Comprehensive LLM extraction performance across task dimensions (Q-2, Q-3).**

| model | family | Q-2-Accuracy | Q-2-Completeness | Q-2-Detail | Q-3-Accuracy | Q-3-Completeness | Q-3-Detail |
|---|---|---|---|---|---|---|---|
| gpt-5-mini | OpenAI | 9.63 | 9.43 | 9.86 | 9.77 | 9.42 | 9.87 |
| gpt-5 | OpenAI | 9.61 | 9.43 | 9.84 | 9.78 | 9.42 | 9.89 |
| gpt-4-1 | OpenAI | 9.73 | 9.42 | 9.85 | 9.84 | 9.28 | 9.75 |
| o4-mini | OpenAI | 9.69 | 9.38 | 9.87 | 9.76 | 9.29 | 9.82 |
| llama3-2 | Local | 9.65 | 9.34 | 9.81 | 9.88 | 9.07 | 9.82 |
| gpt-4o | OpenAI | 9.67 | 9.26 | 9.84 | 9.78 | 8.25 | 9.46 |
| deepseek-r1 | Local | 9.57 | 8.93 | 9.82 | 9.34 | 8.55 | 9.38 |
| llama3 | Local | 9.46 | 9.28 | 9.55 | 9.62 | 8.97 | 9.45 |

Table S4-3: **Comprehensive LLM extraction performance across task dimensions (Q-4, Q-5).**

| model | family | Q-4-Accuracy | Q-4-Completeness | Q-4-Detail | Q-5-Accuracy | Q-5-Completeness | Q-5-Detail |
|---|---|---|---|---|---|---|---|
| gpt-5-mini | OpenAI | 9.88 | 9.43 | 9.88 | 9.72 | 9.08 | 9.88 |
| gpt-5 | OpenAI | 9.88 | 9.35 | 9.81 | 9.73 | 9.05 | 9.83 |
| gpt-4-1 | OpenAI | 9.90 | 9.37 | 9.59 | 9.71 | 8.72 | 9.80 |
| o4-mini | OpenAI | 9.87 | 9.47 | 9.79 | 9.53 | 8.64 | 9.70 |
| llama3-2 | Local | 9.88 | 9.33 | 9.33 | 9.60 | 9.02 | 9.75 |
| gpt-4o | OpenAI | 9.80 | 9.25 | 8.94 | 9.63 | 8.38 | 9.75 |
| deepseek-r1 | Local | 9.36 | 9.39 | 8.48 | 9.76 | 8.54 | 9.80 |
| llama3 | Local | 9.49 | 9.33 | 8.78 | 9.37 | 8.25 | 9.44 |

Table S4-4: **Validation metrics documenting schema compliance and data quality.** Structural validity rates for 7 models evaluated on production datasets. Sample size indicates number of records (title and abstract of a publication) in evaluation; total extractions shows attempted extractions; valid/invalid extractions show results of schema validation; validation issue rate calculated as percentage of invalid extractions. Schema validation performed using predefined JSON schema specifications, testing for: well-formed JSON structure, presence of required fields, correct data types, and adherence to schema-defined field constraints. Models ordered by validation issue rate (ascending). Sample sizes differ by deployment phase: gpt-5 and gpt-4-1 evaluated on full deployment dataset (15,635 records) whereas remaining models on pilot sample (7,000 records).

| model | sample_size | total_extractions | valid_extractions | invalid_extractions | validation_issue_rate |
|---|---|---|---|---|---|
| gpt-5 | 15,635 | 15,606 | 15,606 | 0 | 0.0 |
| gpt-4-1 | 15,635 | 15,628 | 15,626 | 2 | 0.0 |
| o4-mini | 7,000 | 5,369 | 5,366 | 3 | 0.1 |
| llama3-2 | 7,000 | 6,720 | 6,670 | 50 | 0.7 |
| llama3 | 7,000 | 6,901 | 6,416 | 485 | 7.0 |
| deepseek-r1-distilled | 7,000 | 6,184 | 718 | 5,466 | 88.4 |
| gpt-4o | 7,000 | 6,985 | 2 | 6,983 | 100.0 |

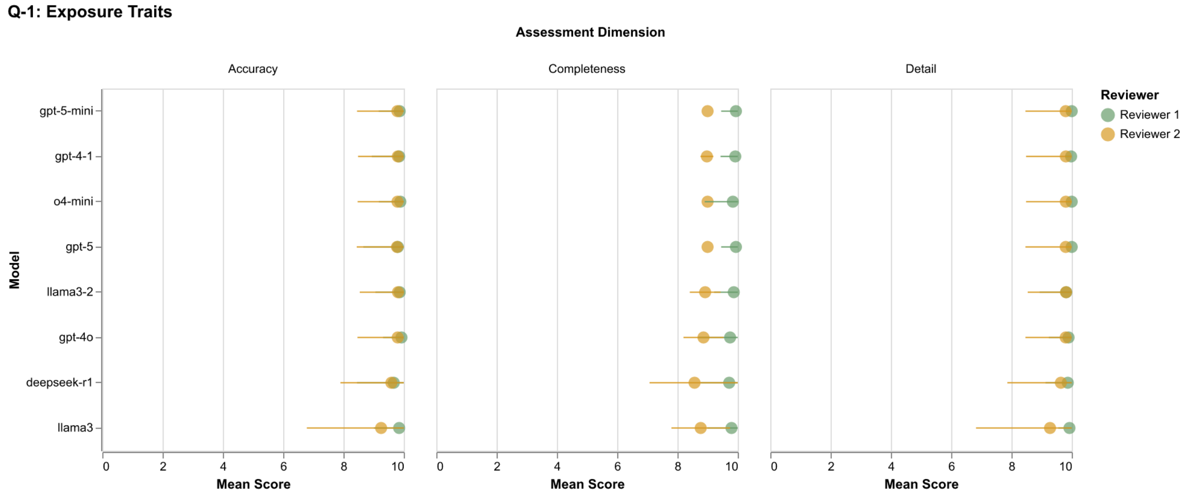


Figure S4-1: **Inter-reviewer comparison of LLM extraction performance for exposure trait information (Q-1).** Mean scores with standard deviation error bars are shown for accuracy, detail, and completeness dimensions across eight LLM models, as assessed by two independent reviewers. Dot plot displays mean assessment scores with error bars representing standard deviation. Green markers indicate Reviewer 1 scores; yellow markers indicate Reviewer 2 scores. Models are ordered by overall mean performance (descending).

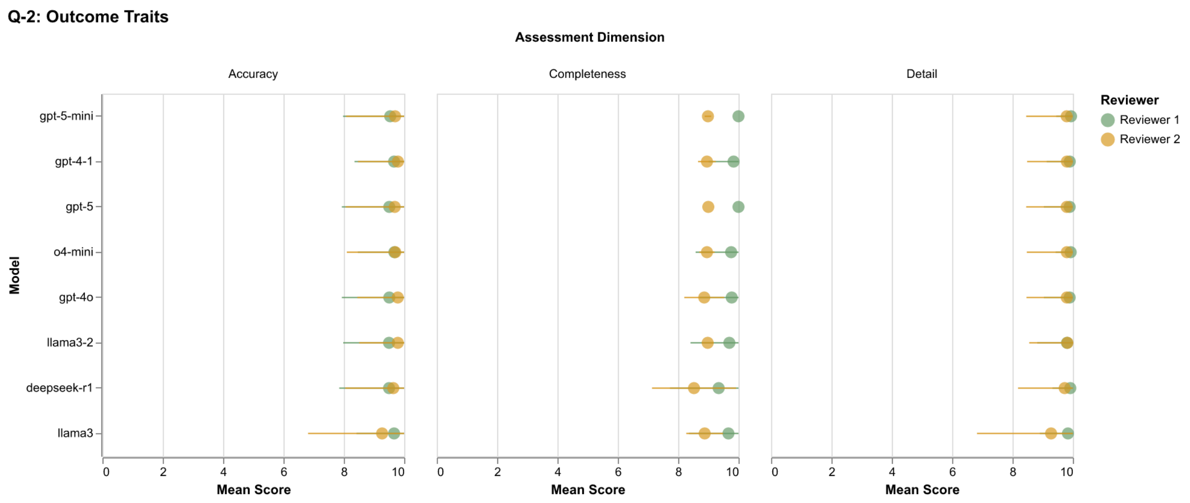


Figure S4-2: **Inter-reviewer comparison of LLM extraction performance for outcome trait information (Q-2).** Mean scores with standard deviation error bars are shown for accuracy, detail, and completeness dimensions across eight LLM models, as assessed by two independent reviewers. Dot plot displays mean assessment scores with error bars representing standard deviation. Green markers indicate Reviewer 1 scores; yellow markers indicate Reviewer 2 scores. Models are ordered by overall mean performance (descending).

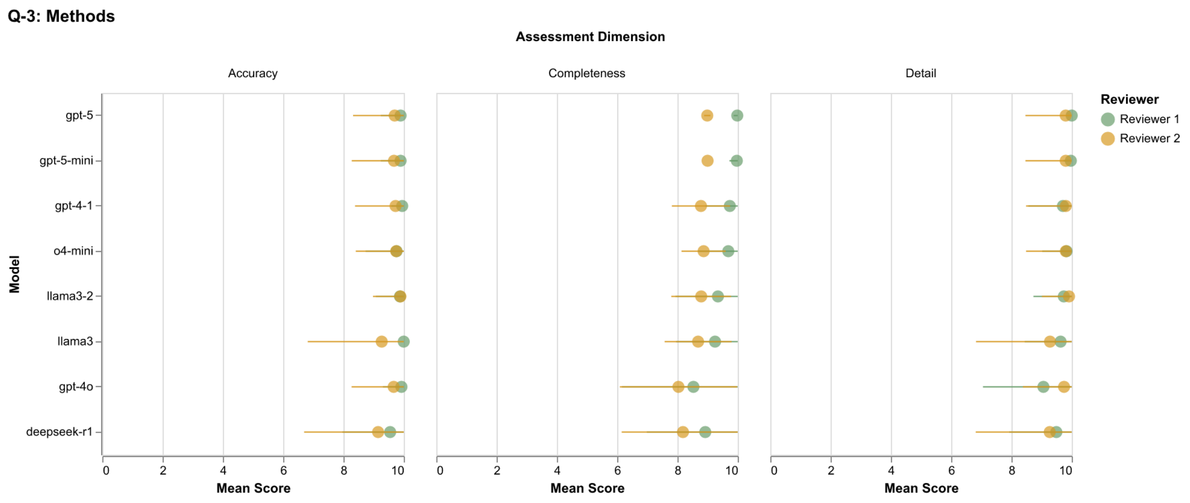


Figure S4-3: **Inter-reviewer comparison of LLM extraction performance for methodological information (Q-3).** Mean scores with standard deviation error bars are shown for accuracy, detail, and completeness dimensions across eight LLM models, as assessed by two independent reviewers. Dot plot displays mean assessment scores with error bars representing standard deviation. Green markers indicate Reviewer 1 scores; yellow markers indicate Reviewer 2 scores. Models are ordered by overall mean performance (descending).

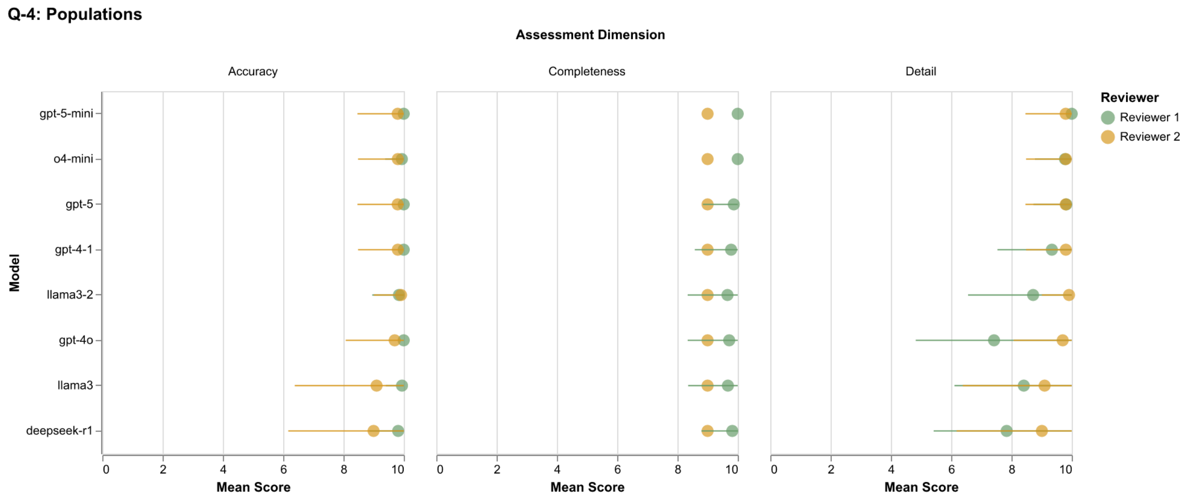


Figure S4-4: **Inter-reviewer comparison of LLM extraction performance for study population information (Q-4).** Mean scores with standard deviation error bars are shown for accuracy, detail, and completeness dimensions across eight LLM models, as assessed by two independent reviewers. Dot plot displays mean assessment scores with error bars representing standard deviation. Green markers indicate Reviewer 1 scores; yellow markers indicate Reviewer 2 scores. Models are ordered by overall mean performance (descending).

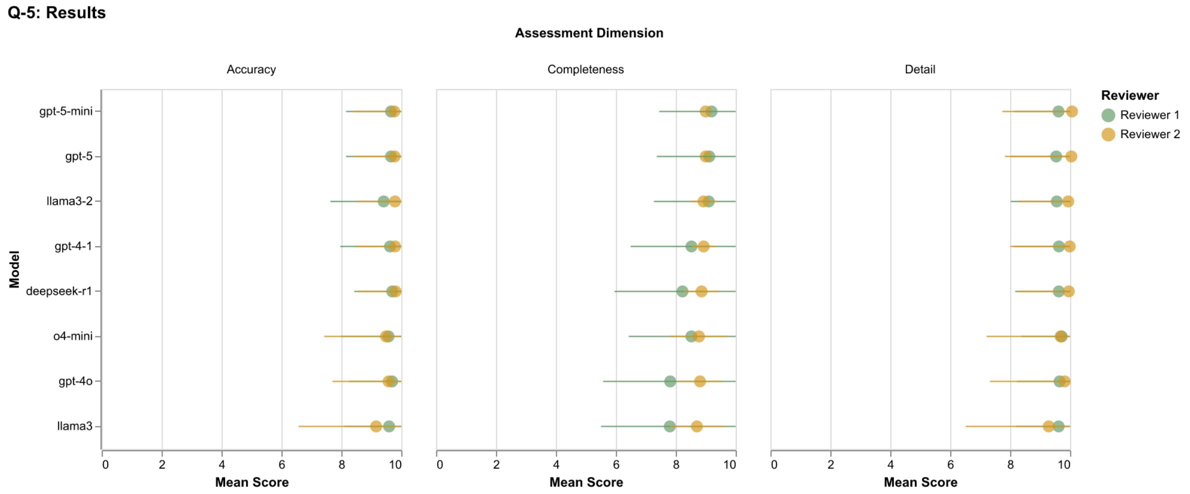


Figure S4-5: **Inter-reviewer comparison of LLM extraction performance for study results (Q-5).** Mean scores with standard deviation error bars are shown for accuracy, detail, and completeness dimensions across eight LLM models, as assessed by two independent reviewers. Dot plot displays mean assessment scores with error bars representing standard deviation. Green markers indicate Reviewer 1 scores; yellow markers indicate Reviewer 2 scores. Models are ordered by overall mean performance (descending).

#### S5 MR-KG summary statistics

Table S5-1: MR-KG Summary Statistics - GPT-4-1 Model

| **Model: GPT-4-1** | |
|---|---|
| Papers processed (PMIDs) | 15,626 |
| Total extraction results | 70,930 |
| Total unique traits | 33,668 |
| Total trait mentions | 80,761 |
| Average results per paper | 4.54 |
| **Trait Profile Similarity** | |
| Total PMID-model combinations | 15,626 |
| Total pairwise comparisons | 156,260 |
| Semantic similarity (mean) | 0.734 |
| Semantic similarity (median) | 0.749 |
| Jaccard similarity (mean) | 0.088 |
| Jaccard similarity (median) | 0.042 |
| Semantic-Jaccard correlation | 0.099 |
| **Evidence Profile Similarity** | |
| Total PMID-model combinations | 1,579 |
| Total pairwise comparisons | 2,066 |
| Direction concordance (mean) | 0.521 |
| Direction concordance (median) | 1.000 |
| Data completeness (mean) | 0.649 |

Table S5-2: MR-KG Summary Statistics - O4-MINI Model

| **Model: O4-MINI** | |
|---|---|
| Papers processed (PMIDs) | 5,366 |
| Total extraction results | 24,046 |
| Total unique traits | 12,567 |
| Total trait mentions | 31,659 |
| Average results per paper | 4.48 |
| **Trait Profile Similarity** | |
| Total PMID-model combinations | 5,366 |
| Total pairwise comparisons | 53,660 |
| Semantic similarity (mean) | 0.701 |
| Semantic similarity (median) | 0.732 |
| Jaccard similarity (mean) | 0.102 |
| Jaccard similarity (median) | 0.062 |
| Semantic-Jaccard correlation | 0.113 |
| **Evidence Profile Similarity** | |
| Total PMID-model combinations | 1,303 |
| Total pairwise comparisons | 1,709 |
| Direction concordance (mean) | 0.409 |
| Direction concordance (median) | 1.000 |
| Data completeness (mean) | 0.755 |

Table S5-3: MR-KG Summary Statistics - LLaMA-3 Model

| **Model: LLaMA-3** | |
|---|---|
| Papers processed (PMIDs) | 6,416 |
| Total extraction results | 26,854 |
| Total unique traits | 10,937 |
| Total trait mentions | 28,027 |
| Average results per paper | 4.19 |
| **Trait Profile Similarity** | |
| Total PMID-model combinations | 6,416 |
| Total pairwise comparisons | 64,160 |
| Semantic similarity (mean) | 0.705 |
| Semantic similarity (median) | 0.732 |
| Jaccard similarity (mean) | 0.102 |
| Jaccard similarity (median) | 0.067 |
| Semantic-Jaccard correlation | 0.253 |
| **Evidence Profile Similarity** | |
| Total PMID-model combinations | 866 |
| Total pairwise comparisons | 608 |
| Direction concordance (mean) | 0.449 |
| Direction concordance (median) | 1.000 |
| Data completeness (mean) | 0.717 |

Table S5-4: MR-KG Summary Statistics - LLaMA-3-2 Model

| **Model: LLaMA-3-2** | |
|---|---|
| Papers processed (PMIDs) | 6,670 |
| Total extraction results | 39,103 |
| Total unique traits | 13,740 |
| Total trait mentions | 33,547 |
| Average results per paper | 5.86 |
| **Trait Profile Similarity** | |
| Total PMID-model combinations | 6,670 |
| Total pairwise comparisons | 66,700 |
| Semantic similarity (mean) | 0.718 |
| Semantic similarity (median) | 0.731 |
| Jaccard similarity (mean) | 0.095 |
| Jaccard similarity (median) | 0.056 |
| Semantic-Jaccard correlation | 0.333 |
| **Evidence Profile Similarity** | |
| Total PMID-model combinations | 709 |
| Total pairwise comparisons | 609 |
| Direction concordance (mean) | 0.311 |
| Direction concordance (median) | 1.000 |
| Data completeness (mean) | 0.596 |

Table S5-5: MR-KG Summary Statistics - Deepseek-R1-Distilled Model

| **Model: Deepseek-R1-Distilled** | |
|---|---|
| Papers processed (PMIDs) | 718 |
| Total extraction results | 3,437 |
| Total unique traits | 2,109 |
| Total trait mentions | 3,197 |
| Average results per paper | 4.79 |
| **Trait Profile Similarity** | |
| Total PMID-model combinations | 718 |
| Total pairwise comparisons | 7,180 |
| Semantic similarity (mean) | 0.588 |
| Semantic similarity (median) | 0.604 |
| Jaccard similarity (mean) | 0.057 |
| Jaccard similarity (median) | 0.000 |
| Semantic-Jaccard correlation | 0.313 |
| **Evidence Profile Similarity** | |
| Total PMID-model combinations | 177 |
| Total pairwise comparisons | 22 |
| Direction concordance (mean) | 0.455 |
| Direction concordance (median) | 1.000 |
| Data completeness (mean) | 0.636 |

### S6 MR-KG individual cases of extracted data

Here we report cases of detailed data for two MR studies on MR-KG, including their abstracts, extracted data, related studies as mapped via trait profile and evidence profile similarities. We used GPT-5 model results for retrieving extracted data and default set of configuration for retrieving related studies. Synthesis on the retrieved results is discussed in Section 4.2.

#### S6.1 PMID: 39836328 [1]

##### Abstract

Study Details
PMID: 39836328

Title: Causal role of ischemic heart disease in ovarian cancer subtypes.

Year: 2024

Journal: Discover oncology

Abstract:

Ischemic heart disease (IHD) may share biological mechanisms with cancer, including ovarian cancer, through pathways such as chronic inflammation and oxidative stress. However, the relationship between IHD and ovarian cancer subtypes remains unclear. This study used Mendelian randomization (MR) to explore potential causal associations.A two-sample MR analysis was conducted using genetic instruments for IHD from large-scale genome-wide association studies (GWAS). The inverse-variance weighted (IVW) method was used as the primary analysis, supported by MR-Egger, weighted median, and MR-PRESSO for sensitivity analyses.No significant association was found between IHD and overall ovarian cancer risk (OR = 0.97, 95% CI 0.92-1.03, P = 0.378). However, IHD was linked to a reduced risk of endometrioid ovarian cancer (OR = 0.86, 95% CI 0.76-0.98, P = 0.027). No associations were observed for serous, mucinous, or clear cell ovarian cancers. Sensitivity analyses confirmed robust findings.IHD may confer a protective effect against endometrioid ovarian cancer but does not influence overall ovarian cancer risk. These findings highlight the need for further research into subtype-specific mechanisms.© 2025. The Author(s).

##### Extracted metadata (JSON)


```
{
  "exposures": [
    {
      "id": 1,
      "trait": "Ischemic heart disease (IHD)",
      "category": "disease of the circulatory system",
      "linked_index": 2188
    }
  ],
  "outcomes": [
    {
      "id": 1,
      "trait": "Ovarian cancer (overall risk)",
      "category": "neoplasm",
      "linked_index": 48590
    },
    {
      "id": 2,
      "trait": "Endometrioid ovarian cancer",
      "category": "neoplasm",
      "linked_index": 2104
    },
    {
      "id": 3,
      "trait": "Serous ovarian cancer",
      "category": "neoplasm",
      "linked_index": 28981
    },
    {
      "id": 4,
      "trait": "Mucinous ovarian cancer",
      "category": "neoplasm",
      "linked_index": 29675
    },
    {
      "id": 5,
      "trait": "Clear cell ovarian cancer",
      "category": "neoplasm",
      "linked_index": 2103
    }
  ],
  "methods": [
    "two-sample mendelian randomization",
    "sensitivity analysis",
    "Other: inverse-variance weighted (IVW)",
    "Other: MR-Egger",
    "Other: weighted median",
    "Other: MR-PRESSO"
  ],
  "population": []
}
```


##### Extracted results (JSON)

```
[
  {
    "exposure": "Ischemic heart disease (IHD)",
    "outcome": "Overall ovarian cancer risk",
    "beta": null,
    "units": null,
    "hazard ratio": null,
    "odds ratio": 0.97,
    "95% CI": [0.92, 1.03],
    "SE": null,
    "P-value": 0.378,
    "direction": "no association"
  },
  {
    "exposure": "Ischemic heart disease (IHD)",
    "outcome": "Endometrioid ovarian cancer",
    "beta": null,
    "units": null,
    "hazard ratio": null,
    "odds ratio": 0.86,
    "95% CI": [0.76, 0.98],
    "SE": null,
    "P-value": 0.027,
    "direction": "decreases"
  },
  {
    "exposure": "Ischemic heart disease (IHD)",
    "outcome": "Serous ovarian cancer",
    "beta": null,
    "units": null,
    "hazard ratio": null,
    "odds ratio": null,
    "95% CI": [null, null],
    "SE": null,
    "P-value": null,
    "direction": "no association"
  },
  {
    "exposure": "Ischemic heart disease (IHD)",
    "outcome": "Mucinous ovarian cancer",
    "beta": null,
    "units": null,
    "hazard ratio": null,
    "odds ratio": null,
    "95% CI": [null, null],
    "SE": null,
    "P-value": null,
    "direction": "no association"
  },
  {
    "exposure": "Ischemic heart disease (IHD)",
    "outcome": "Clear cell ovarian cancer",
    "beta": null,
    "units": null,
    "hazard ratio": null,
    "odds ratio": null,
    "95% CI": [null, null],
    "SE": null,
    "P-value": null,
```

```
    "direction": "no association"
  }
]
```


**Trait profile similarity results (JSON)**


```
{
  "query_pmid": "39836328",
  "query_model": "gpt-5",
  "query_title": "Causal role of ischemic heart disease in ovarian cancer subtypes.",
  "query_trait_count": 6,
  "similar_studies": [
    {
      "pmid": "38090480",
      "title": "Mendelian randomization analysis to elucidate the causal relationship
between small molecule metabolites and ovarian cancer risk.",
      "trait_profile_similarity": 0.8752651611963908,
      "trait_jaccard_similarity": 0.25,
      "trait_count": 9,
      "involved_traits": [
        { "trait_index": 2103, "trait_label": "Clear cell ovarian cancer" },
        { "trait_index": 2104, "trait_label": "Endometrioid ovarian cancer" },
        { "trait_index": 43538, "trait_label": "Hexadecenoylcarnitine" },
        {
          "trait_index": 10484,
          "trait_label": "High-grade serous ovarian cancer"
        },
        { "trait_index": 43539, "trait_label": "Methionine sulfoxide" },
        {
          "trait_index": 48590,
          "trait_label": "Ovarian cancer (overall risk)"
        },
        {
          "trait_index": 72672,
          "trait_label": "Small molecule metabolites (53 metabolites from GWAS)"
        },
        { "trait_index": 43540, "trait_label": "Tetradecanoyl carnitine" },
        { "trait_index": 1227, "trait_label": "Tryptophan" }
      ]
    },
    {
      "pmid": "34669940",
      "title": "Causes and Consequences of Polycystic Ovary Syndrome: Insights From
Mendelian Randomization.",
      "trait_profile_similarity": 0.8604791462421417,
      "trait_jaccard_similarity": 0.05555555555555555,
      "trait_count": 14,
      "involved_traits": [
        { "trait_index": 363, "trait_label": "Coronary heart disease" },
        { "trait_index": 90, "trait_label": "Depression" },
        { "trait_index": 2104, "trait_label": "Endometrioid ovarian cancer" },
        {
          "trait_index": 3567,
          "trait_label": "Estrogen receptor-positive breast cancer"
        },
        { "trait_index": 198, "trait_label": "Fasting insulin" },
        { "trait_index": 22808, "trait_label": "Male-pattern balding" },
        { "trait_index": 22807, "trait_label": "Menopause timing" },
        { "trait_index": 547, "trait_label": "Obesity" },
```

```
        {
          "trait_index": 1296,
          "trait_label": "Polycystic ovary syndrome (PCOS)"
        },
        {
          "trait_index": 37430,
          "trait_label": "Serum sex hormone-binding globulin concentrations"
        },
        { "trait_index": 185, "trait_label": "Stroke" },
        { "trait_index": 8716, "trait_label": "Testosterone levels" },
        { "trait_index": 246, "trait_label": "Type 2 diabetes" }
      ]
    },
    {
      "pmid": "28241208",
      "title": "Association Between Telomere Length and Risk of Cancer and Non-Neoplastic
Diseases: A Mendelian Randomization Study.",
      "trait_profile_similarity": 0.8465254306793213,
      "trait_jaccard_similarity": 0,
      "trait_count": 19,
      "involved_traits": [
        { "trait_index": 4013, "trait_label": "Abdominal aortic aneurysm" },
        { "trait_index": 2825, "trait_label": "Autoimmune diseases" },
        { "trait_index": 1078, "trait_label": "Bladder cancer" },
        { "trait_index": 1391, "trait_label": "Celiac disease" },
        { "trait_index": 363, "trait_label": "Coronary heart disease" },
        { "trait_index": 2400, "trait_label": "Diabetes mellitus" },
        { "trait_index": 531, "trait_label": "Endometrial cancer" },
        { "trait_index": 1394, "trait_label": "Glioma" },
        { "trait_index": 49754, "trait_label": "Inflammatory diseases" },
        { "trait_index": 3743, "trait_label": "Interstitial lung disease" },
        { "trait_index": 378, "trait_label": "Kidney cancer" },
        { "trait_index": 100, "trait_label": "Lung adenocarcinoma" },
        { "trait_index": 1464, "trait_label": "Melanoma" },
        { "trait_index": 8064, "trait_label": "Neuroblastoma" },
        {
          "trait_index": 53008,
          "trait_label": "Other non-neoplastic diseases"
        },
        { "trait_index": 4678, "trait_label": "Psychiatric disorders" },
        {
          "trait_index": 53007,
          "trait_label": "Serous low-malignant-potential ovarian cancer"
        },
        { "trait_index": 1554, "trait_label": "Telomere length" },
        { "trait_index": 781, "trait_label": "Testicular cancer" }
      ]
    },
    {
      "pmid": "39519507",
      "title": "Using Genetics to Assess the Role of Acetate in Ischemic Heart Disease,
Diabetes, and Sex-Hormone-Related Cancers: A Mendelian Randomization Study.",
      "trait_profile_similarity": 0.8276673754056295,
      "trait_jaccard_similarity": 0,
      "trait_count": 8,
      "involved_traits": [
        { "trait_index": 27007, "trait_label": "Acetate" },
        { "trait_index": 176, "trait_label": "Breast cancer" },
        { "trait_index": 268, "trait_label": "Colorectal cancer" },
```

```
        { "trait_index": 506, "trait_label": "Diabetes" },
        { "trait_index": 531, "trait_label": "Endometrial cancer" },
        { "trait_index": 380, "trait_label": "Ischemic heart disease" },
        { "trait_index": 596, "trait_label": "Ovarian cancer" },
        { "trait_index": 365, "trait_label": "Prostate cancer" }
      ]
    },
    {
      "pmid": "35492879",
      "title": "A multi-level investigation of the genetic relationship between
endometriosis and ovarian cancer histotypes.",
      "trait_profile_similarity": 0.824830504755179,
      "trait_jaccard_similarity": 0.2222222222222222,
      "trait_count": 5,
      "involved_traits": [
        { "trait_index": 2103, "trait_label": "Clear cell ovarian cancer" },
        { "trait_index": 2104, "trait_label": "Endometrioid ovarian cancer" },
        { "trait_index": 103, "trait_label": "Endometriosis" },
        {
          "trait_index": 52027,
          "trait_label": "Epithelial ovarian cancer (overall)"
        },
        {
          "trait_index": 10484,
          "trait_label": "High-grade serous ovarian cancer"
        }
      ]
    },
    {
      "pmid": "34601599",
      "title": "Serum Estradiol and 20 Site-Specific Cancers in Women: Mendelian
Randomization Study.",
      "trait_profile_similarity": 0.8221626927455267,
      "trait_jaccard_similarity": 0.07142857142857142,
      "trait_count": 9,
      "involved_traits": [
        { "trait_index": 75067, "trait_label": "Any cancer (women)" },
        { "trait_index": 20484, "trait_label": "Breast cancer (overall)" },
        { "trait_index": 46185, "trait_label": "Endometrial cancer (overall)" },
        {
          "trait_index": 2843,
          "trait_label": "Endometrioid endometrial cancer"
        },
        { "trait_index": 2104, "trait_label": "Endometrioid ovarian cancer" },
        {
          "trait_index": 1653,
          "trait_label": "Estrogen receptor (ER)-positive breast cancer"
        },
        { "trait_index": 36809, "trait_label": "Ovarian cancer (overall)" },
        {
          "trait_index": 75066,
          "trait_label": "Serum endogenous 17 -estradiol (E2) levels"
        },
        { "trait_index": 46297, "trait_label": "Stomach (gastric) cancer" }
      ]
    },
    {
      "pmid": "36937699",
```

```
      "title": "Causal effects of physical activity on the risk of overall ovarian cancer: A Mendelian randomization study.",
      "trait_profile_similarity": 0.8221626927455267,
      "trait_jaccard_similarity": 0.09090909090909091,
      "trait_count": 6,
      "involved_traits": [
        {
          "trait_index": 20952,
          "trait_label": "Accelerometer-measured average acceleration"
        },
        {
          "trait_index": 20954,
          "trait_label": "Accelerometer-measured overall accelerations"
        },
        {
          "trait_index": 20953,
          "trait_label": "Accelerometer-measured overall activity"
        },
        { "trait_index": 2104, "trait_label": "Endometrioid ovarian cancer" },
        { "trait_index": 36809, "trait_label": "Ovarian cancer (overall)" },
        {
          "trait_index": 12491,
          "trait_label": "Self-reported moderate-to-vigorous physical activity"
        }
      ]
    },
    {
      "pmid": "35094873",
      "title": "Mendelian randomization analyses for PCOS: evidence, opportunities, and challenges.",
      "trait_profile_similarity": 0.8214789927005768,
      "trait_jaccard_similarity": 0,
      "trait_count": 18,
      "involved_traits": [
        { "trait_index": 368, "trait_label": "Adiposity" },
        {
          "trait_index": 5774,
          "trait_label": "Anti-Müllerian hormone (AMH) levels"
        },
        { "trait_index": 3153, "trait_label": "Anxiety disorder (AD)" },
        { "trait_index": 3154, "trait_label": "Bipolar disorder (BIP)" },
        { "trait_index": 176, "trait_label": "Breast cancer" },
        { "trait_index": 278, "trait_label": "Coronary heart disease (CHD)" },
        { "trait_index": 90, "trait_label": "Depression" },
        { "trait_index": 947, "trait_label": "Forced vital capacity (FVC)" },
        { "trait_index": 5259, "trait_label": "Insulin resistance (IR)" },
        { "trait_index": 9549, "trait_label": "Menopause age" },
        {
          "trait_index": 4575,
          "trait_label": "Obsessive-compulsive disorder (OCD)"
        },
        { "trait_index": 1805, "trait_label": "Offspring birth weight" },
        { "trait_index": 596, "trait_label": "Ovarian cancer" },
        {
          "trait_index": 1296,
          "trait_label": "Polycystic ovary syndrome (PCOS)"
        },
        { "trait_index": 595, "trait_label": "Schizophrenia (SCZ)" },
        {
```

```
          "trait_index": 2475,
          "trait_label": "Sex hormone-binding globulin (SHBG) levels"
        },
        { "trait_index": 185, "trait_label": "Stroke" },
        { "trait_index": 197, "trait_label": "Type 2 diabetes mellitus (T2DM)" }
      ]
    },
    {
      "pmid": "34671129",
      "title": "Identifying causality, genetic correlation, priority and pathways of
large-scale complex exposures of breast and ovarian cancers.",
      "trait_profile_similarity": 0.8213934948047003,
      "trait_jaccard_similarity": 0,
      "trait_count": 25,
      "involved_traits": [
        { "trait_index": 362, "trait_label": "Adiponectin" },
        { "trait_index": 1358, "trait_label": "Age at menopause" },
        { "trait_index": 1009, "trait_label": "Basal metabolic rate" },
        { "trait_index": 93, "trait_label": "Birth weight" },
        { "trait_index": 5232, "trait_label": "Body fat mass (BFM)" },
        { "trait_index": 97, "trait_label": "Body fat percentage" },
        { "trait_index": 6940, "trait_label": "Body fat-free mass" },
        { "trait_index": 63, "trait_label": "Body mass index (BMI)" },
        { "trait_index": 20484, "trait_label": "Breast cancer (overall)" },
        {
          "trait_index": 62781,
          "trait_label": "Breast cancer, oestrogen receptor-negative (ER-)"
        },
        {
          "trait_index": 62780,
          "trait_label": "Breast cancer, oestrogen receptor-positive (ER+)"
        },
        { "trait_index": 763, "trait_label": "Childhood obesity" },
        {
          "trait_index": 7518,
          "trait_label": "Clear cell ovarian cancer (CCOC)"
        },
        {
          "trait_index": 6938,
          "trait_label": "Comparative body size at age 10 (CBS-10)"
        },
        { "trait_index": 3281, "trait_label": "Education attainment" },
        {
          "trait_index": 48922,
          "trait_label": "Endometrioid ovarian cancer (EOC)"
        },
        {
          "trait_index": 7521,
          "trait_label": "High-grade serous ovarian cancer (HGSOC)"
        },
        {
          "trait_index": 7520,
          "trait_label": "Invasive mucinous ovarian cancer (IMOC)"
        },
        { "trait_index": 1298, "trait_label": "Linoleic acid" },
        {
          "trait_index": 15559,
          "trait_label": "Low-grade serous ovarian cancer"
        },
```

```
        { "trait_index": 725, "trait_label": "Omega-6 fatty acids" },
        {
          "trait_index": 62779,
          "trait_label": "Omega-6:omega-3 fatty acids ratio"
        },
        { "trait_index": 36809, "trait_label": "Ovarian cancer (overall)" },
        { "trait_index": 22, "trait_label": "Schizophrenia" },
        { "trait_index": 681, "trait_label": "Waist circumference (WC)" }
      ]
    },
    {
      "pmid": "38822303",
      "title": "Causal relationship between lipid-lowering drugs and ovarian cancer, cervical cancer: a drug target mendelian randomization study.",
      "trait_profile_similarity": 0.8082764943440756,
      "trait_jaccard_similarity": 0,
      "trait_count": 5,
      "involved_traits": [
        { "trait_index": 1257, "trait_label": "Cervical cancer" },
        { "trait_index": 363, "trait_label": "Coronary heart disease" },
        { "trait_index": 993, "trait_label": "HMGCR inhibitors" },
        { "trait_index": 596, "trait_label": "Ovarian cancer" },
        { "trait_index": 994, "trait_label": "PCSK9 inhibitors" }
      ]
    }
  ]
}
```


**Evidence profile similarity results (JSON)**


```
{
  "query_pmid": "39836328",
  "query_model": "gpt-5",
  "query_title": "Causal role of ischemic heart disease in ovarian cancer subtypes.",
  "query_result_count": 5,
  "similar_studies": [
    {
      "pmid": "39719456",
      "title": "Association of cardiovascular disease on cancer: observational and mendelian randomization analyses.",
      "direction_concordance": 1,
      "matched_pairs": 1,
      "match_type_exact": false,
      "match_type_fuzzy": true,
      "match_type_efo": false,
      "matched_evidence_pairs": [
        {
          "query_exposure": "Ischemic heart disease (IHD)",
          "query_outcome": "Endometrioid ovarian cancer",
          "query_direction": "decreases",
          "similar_exposure": "Coronary heart disease",
          "similar_outcome": "Breast cancer",
          "similar_direction": "decreases",
          "match_type": "fuzzy"
        }
      ]
    }
  ]
}
```

##### S6.2 PMID: 40325806 [2]

###### Abstract

Study Details
PMID: 40325806

Title: Unveiling the Role of Immune Cells and Plasma Metabolites in Breast Cancer Risk: A Mendelian Randomization and Mediation Analysis.

Year: 2024

Journal: Current pharmaceutical biotechnology

Abstract:

Breast cancer (BC) is the most common cancer worldwide, yet identifying effective therapeutic targets continues to pose challenges. To understand the biological mechanisms driving BC and uncover potential therapeutic strategies, we performed a comprehensive Mendelian randomization (MR) analysis. The objective of this study is to investigate the causal relationships between immune cell phenotypes and BC risk, with a focus on identifying intermediary metabolites involved in these processes.We conducted MR analysis using genome-wide association study (GWAS) data from publicly available databases, focusing on 731 immune cell traits (n = 3757), 1400 plasma metabolites (n = 8299), and breast cancer (n case = 6188, n control = 182,678). We used a two-step MR approach to examine the potential intermediary role of plasma metabolites in the immune cell-BC relationship. The research employs the inverse variance weighting (IVW) method as its primary approach. To assess potential sources of bias, such as horizontal pleiotropy and heterogeneity, we employed the MR Egger intercept test and the Cochran's Q test, respectively. These rigorous analytical approaches ensured the robustness of our findings. Furthermore, Bayesian colocalization was employed to enhance the accuracy of causal inference and reduce false positives.The IVW method of reverse MR analysis revealed a causal relationship between 20 immune cell traits and BC. Notably, our analysis identified three plasma metabolites as potential mediators in the causal link between immune cells and BC. In particular, vanillic acid glycine partially mediated the relationship between CD28+ CD45RA- CD8dim %CD8dim cells and BC. This suggests that vanillic acid glycine may act as a molecular link, modulating immune cell functions that contribute to BC. Additionally, the pyruvate to N-acetylneuraminate ratio exhibited negative mediation effects involving IgD- CD38dim AC, HLA DR on DC, CD8dim NKT %T cells, and CCR2 on monocytes. The negative mediation implies that this metabolite might inhibit certain immune cell behaviors that would otherwise promote cancer progression. Furthermore, cysteinylglycine disulfide partially mediated the relationship between CD8 on TD CD8br cells and BC, suggesting its involvement in the modulation of CD8 +T cell responses in BC. Key immune cell phenotypes, such as CCR2 on monocytes (odds ratio, ORIVW = 0.969; 95% CI: 0.943-0.996; p = 0.027), IgD- CD38dim AC cells (ORIVW = 1.100; 95% CI: 1.023-1.183; p = 0.009), were highlighted for their significant roles in BC progression.This study shows that vanillic acid glycine levels, the pyruvate to N-acetylneuraminate ratio, and cysteinylglycine disulfide levels can act as mediators in reducing breast cancer risk. These findings support the idea that metabolic pathways and metabolites are crucial in cancer progression, offering insights into potential biomarkers for early detection, treatment assessment, and recurrence prediction, thus paving the way for more personalized clinical approaches.Copyright© Bentham Science Publishers; For any queries, please email at.

###### Extracted metadata (JSON)


```
[
  {
    "exposure": "CCR2 on monocytes",
    "outcome": "Breast cancer",
```

```
    "beta": null,
    "units": null,
    "hazard ratio": null,
    "odds ratio": 0.969,
    "95% CI": [0.943, 0.996],
    "SE": null,
    "P-value": 0.027,
    "direction": "decreases"
  },
  {
    "exposure": "IgD- CD38dim AC cells",
    "outcome": "Breast cancer",
    "beta": null,
    "units": null,
    "hazard ratio": null,
    "odds ratio": 1.1,
    "95% CI": [1.023, 1.183],
    "SE": null,
    "P-value": 0.009,
    "direction": "increases"
  },
  {
    "exposure": "Vanillic acid glycine",
    "outcome": "Breast cancer",
    "beta": null,
    "units": null,
    "hazard ratio": null,
    "odds ratio": null,
    "95% CI": [null, null],
    "SE": null,
    "P-value": null,
    "direction": "decreases"
  },
  {
    "exposure": "Pyruvate to N-acetylneuraminate ratio",
    "outcome": "Breast cancer",
    "beta": null,
    "units": null,
    "hazard ratio": null,
    "odds ratio": null,
    "95% CI": [null, null],
    "SE": null,
    "P-value": null,
    "direction": "decreases"
  },
  {
    "exposure": "Cysteinylglycine disulfide",
    "outcome": "Breast cancer",
    "beta": null,
    "units": null,
    "hazard ratio": null,
    "odds ratio": null,
    "95% CI": [null, null],
    "SE": null,
    "P-value": null,
    "direction": "decreases"
  }
]
```

#### Extracted results (JSON)


```
{
  "exposures": [
    {
      "id": 1,
      "trait": "Immune cell traits (731 phenotypes)",
      "category": "molecular",
      "linked_index": 51184
    },
    {
      "id": 2,
      "trait": "Vanillic acid glycine",
      "category": "molecular",
      "linked_index": 51185
    },
    {
      "id": 3,
      "trait": "Pyruvate to N-acetylneuraminate ratio",
      "category": "molecular",
      "linked_index": 51186
    },
    {
      "id": 4,
      "trait": "Cysteinylglycine disulfide",
      "category": "molecular",
      "linked_index": 51187
    },
    {
      "id": 5,
      "trait": "CCR2 on monocytes",
      "category": "molecular",
      "linked_index": 18347
    },
    {
      "id": 6,
      "trait": "IgD- CD38dim AC cells",
      "category": "molecular",
      "linked_index": 51188
    },
    {
      "id": 7,
      "trait": "HLA-DR on dendritic cells (DC)",
      "category": "molecular",
      "linked_index": 51189
    },
    {
      "id": 8,
      "trait": "CD8dim NKT %T cells",
      "category": "molecular",
      "linked_index": 51190
    },
    {
      "id": 9,
      "trait": "CD8 on TD CD8bright cells",
      "category": "molecular",
      "linked_index": 51191
    },
    {
```

```
      "id": 10,
      "trait": "CD28+ CD45RA- CD8dim %CD8dim cells",
      "category": "molecular",
      "linked_index": 51192
    }
  ],
  "outcomes": [
    {
      "id": 1,
      "trait": "Breast cancer",
      "category": "neoplasm",
      "linked_index": 176
    }
  ],
  "methods": [
    "reverse mendelian randomization",
    "colocalization",
    "sensitivity analysis",
    { "Other": "inverse variance weighting (IVW)" },
    { "Other": "two-step Mendelian randomization" },
    { "Other": "MR Egger intercept test" },
    { "Other": "Cochran's Q test" }
  ],
  "population": [
    "GWAS participants from publicly available databases: immune cell traits n=3,757",
    "GWAS participants from publicly available databases: plasma metabolites n=8,299",
    "Breast cancer GWAS: 6,188 cases and 182,678 controls",
    "Ancestry not specified"
  ]
}
```


##### Trait profile similarity results (JSON)


```
{
  "query_pmid": "40325806",
  "query_model": "gpt-5",
  "query_title": "Unveiling the Role of Immune Cells and Plasma Metabolites in Breast
Cancer Risk: A Mendelian Randomization and Mediation Analysis.",
  "query_trait_count": 11,
  "similar_studies": [
    {
      "pmid": "39884473",
      "title": "Causal association and potential mediator between lung cancer and
tuberculosis: A Mendelian randomization study.",
      "trait_profile_similarity": 0.7046116346662695,
      "trait_jaccard_similarity": 0,
      "trait_count": 14,
      "involved_traits": [
        {
          "trait_index": 59269,
          "trait_label": "CD4 on CD39+ resting regulatory T cells"
        },
        { "trait_index": 59271, "trait_label": "Dopamine 4-sulfate" },
        { "trait_index": 59270, "trait_label": "Ethylparaben sulfate" },
        {
          "trait_index": 59267,
          "trait_label": "Immunophenotypes (various, 731 traits)"
        },
        { "trait_index": 3505, "trait_label": "Lung cancer (LC)" },
```

```
        { "trait_index": 48568, "trait_label": "N-formylmethionine" },
        { "trait_index": 59273, "trait_label": "N6,N6,N6-trimethyllysine" },
        { "trait_index": 57943, "trait_label": "Pantoate" },
        { "trait_index": 59272, "trait_label": "Pentose acid" },
        {
          "trait_index": 59268,
          "trait_label": "Plasma metabolites (various, 1400 traits)"
        },
        { "trait_index": 59274, "trait_label": "Tryptophan to pyruvate ratio" },
        { "trait_index": 1913, "trait_label": "Tuberculosis (TB)" }
      ]
    },
    {
      "pmid": "39346920",
      "title": "Causal effects and metabolites mediators between immune cell and risk of colorectal cancer: a Mendelian randomization study.",
      "trait_profile_similarity": 0.7020849504254081,
      "trait_jaccard_similarity": 0,
      "trait_count": 10,
      "involved_traits": [
        { "trait_index": 62042, "trait_label": "AMP to aspartate ratio" },
        { "trait_index": 47381, "trait_label": "BAFF-R on B cells" },
        { "trait_index": 62039, "trait_label": "CD14+ CD16- monocytes" },
        { "trait_index": 48, "trait_label": "Colorectal cancer (CRC)" },
        { "trait_index": 62040, "trait_label": "IgD+ CD38bright %B cells" },
        { "trait_index": 62044, "trait_label": "Iminodiacetate (IDA)" },
        {
          "trait_index": 62041,
          "trait_label": "Metabolites (blood metabolome)"
        },
        {
          "trait_index": 62038,
          "trait_label": "Myeloid dendritic cells (%DC)"
        },
        {
          "trait_index": 62037,
          "trait_label": "Peripheral immune cell types (genetically inferred)"
        },
        {
          "trait_index": 62043,
          "trait_label": "Retinol (Vitamin A) to linoleoyl-arachidonoyl-glycerol (18:2/20:4) ratio"
        }
      ]
    },
    {
      "pmid": "39465812",
      "title": "Mediation effect of plasma metabolites on the relationship between immune cells and the risk of prostatitis: A study by bidirectional 2-sample and Bayesian-weighted Mendelian randomization.",
      "trait_profile_similarity": 0.6864624551751397,
      "trait_jaccard_similarity": 0,
      "trait_count": 9,
      "involved_traits": [
        {
          "trait_index": 52216,
          "trait_label": "CD14-CD16+ monocyte absolute count"
        },
        {
```

```
          "trait_index": 52215,
          "trait_label": "CD3 on CD39+ activated regulatory T cells (activated Treg)"
        },
        {
          "trait_index": 52221,
          "trait_label": "Glutamine degradant (plasma metabolite)"
        },
        { "trait_index": 52217, "trait_label": "HLA-DR+ CD4+ percent T cells" },
        {
          "trait_index": 52219,
          "trait_label": "Histidine betaine (hercynine)"
        },
        {
          "trait_index": 52214,
          "trait_label": "Immunophenotypes (immune cell subsets; 731 traits analyzed)"
        },
        { "trait_index": 52220, "trait_label": "Proline-to-glutamate ratio" },
        { "trait_index": 3841, "trait_label": "Prostatitis" },
        { "trait_index": 52218, "trait_label": "X-24344 (plasma metabolite)" }
      ]
    },
    {
      "pmid": "39403270",
      "title": "Genetic causality of lipidomic and immune cell profiles in ischemic stroke.",
      "trait_profile_similarity": 0.6796094233339484,
      "trait_jaccard_similarity": 0,
      "trait_count": 16,
      "involved_traits": [
        { "trait_index": 74752, "trait_label": "CCR2 on granulocytes" },
        {
          "trait_index": 74753,
          "trait_label": "CD11c on CD62L+ myeloid dendritic cells"
        },
        {
          "trait_index": 74755,
          "trait_label": "CD4 on activated & secreting CD4 regulatory T cells"
        },
        {
          "trait_index": 74754,
          "trait_label": "CD4 on activated CD4 regulatory T cells"
        },
        { "trait_index": 74756, "trait_label": "Cardioembolic stroke (CS)" },
        { "trait_index": 51986, "trait_label": "FSC-A on granulocytes" },
        {
          "trait_index": 48206,
          "trait_label": "Immune cell phenotypes (731 traits)"
        },
        { "trait_index": 74209, "trait_label": "Ischemic stroke (overall)" },
        { "trait_index": 612, "trait_label": "Large artery stroke (LAS)" },
        {
          "trait_index": 53697,
          "trait_label": "Lipid species (179 lipidomic traits)"
        },
        { "trait_index": 16453, "trait_label": "Phosphatidylcholines" },
        { "trait_index": 29834, "trait_label": "Phosphatidylethanolamines" },
        { "trait_index": 29835, "trait_label": "Phosphatidylinositols" },
        { "trait_index": 611, "trait_label": "Small vessel stroke (SVS)" },
        { "trait_index": 8258, "trait_label": "Sphingomyelin" },
```

```
        { "trait_index": 57963, "trait_label": "Sterol esters" }
      ]
    },
    {
      "pmid": "38826800",
      "title": "Causal effects and metabolites mediators between immune cell and risk of
breast cancer: a Mendelian randomization study.",
      "trait_profile_similarity": 0.6736521179025824,
      "trait_jaccard_similarity": 0.05263157894736842,
      "trait_count": 9,
      "involved_traits": [
        { "trait_index": 65680, "trait_label": "Blood immune cell levels" },
        { "trait_index": 176, "trait_label": "Breast cancer" },
        { "trait_index": 2710, "trait_label": "CD14+ CD16+ monocytes" },
        { "trait_index": 37798, "trait_label": "CD24+ CD27+ B cells" },
        { "trait_index": 37800, "trait_label": "Glycerate" },
        { "trait_index": 65681, "trait_label": "Glycerate levels" },
        { "trait_index": 37799, "trait_label": "IgD- CD38+ B cells" },
        { "trait_index": 37801, "trait_label": "Succinoyltaurine" },
        { "trait_index": 65682, "trait_label": "Succinoyltaurine levels" }
      ]
    },
    {
      "pmid": "38327747",
      "title": "A Mendelian analysis of the relationships between immune cells and breast
cancer.",
      "trait_profile_similarity": 0.6629185947504911,
      "trait_jaccard_similarity": 0,
      "trait_count": 11,
      "involved_traits": [
        { "trait_index": 7971, "trait_label": "BAFF-R on IgD+ CD38- unsw mem" },
        { "trait_index": 20484, "trait_label": "Breast cancer (overall)" },
        { "trait_index": 7973, "trait_label": "CD19 on IgD- CD38br" },
        { "trait_index": 7974, "trait_label": "CD25 on IgD+ CD38dim" },
        { "trait_index": 7972, "trait_label": "CD27 on PB/PC" },
        { "trait_index": 7969, "trait_label": "CD45RA- CD4+ %CD4+" },
        { "trait_index": 7970, "trait_label": "CD8dim %T cell" },
        {
          "trait_index": 1855,
          "trait_label": "CX3CR1 on CD14+ CD16- monocyte"
        },
        { "trait_index": 115, "trait_label": "ER-negative breast cancer" },
        { "trait_index": 10969, "trait_label": "ER-positive breast cancer" },
        {
          "trait_index": 61420,
          "trait_label": "Immune cell traits (731 immune cell phenotypes)"
        }
      ]
    },
    {
      "pmid": "39698561",
      "title": "Causal relationship between immune cells, inflammatory cytokines,
metabolites, and erectile dysfunction: a two-sample Mendelian randomization study.",
      "trait_profile_similarity": 0.662593660029498,
      "trait_jaccard_similarity": 0,
      "trait_count": 16,
      "involved_traits": [
        {
          "trait_index": 71135,
```

```
          "trait_label": "4-methyl-2-oxopentanoate to 3-methyl-2-oxobutyrate ratio"
        },
        {
          "trait_index": 71136,
          "trait_label": "Alpha-ketoglutarate to kynurenine ratio"
        },
        {
          "trait_index": 71133,
          "trait_label": "Aspartate to N-acetylglucosamine to N-acetylgalactosamine ratio"
        },
        { "trait_index": 71128, "trait_label": "CD19 on IgD- CD38+ B cells" },
        { "trait_index": 71130, "trait_label": "CD25 on IgD+ CD24- B cells" },
        { "trait_index": 42667, "trait_label": "CD25 on IgD+ CD38dim B cells" },
        {
          "trait_index": 71129,
          "trait_label": "CD4 on terminally differentiated CD4+ T cells"
        },
        {
          "trait_index": 71134,
          "trait_label": "Cholesterol to taurocholate ratio"
        },
        {
          "trait_index": 71127,
          "trait_label": "Circulating metabolites (1,400 traits from GWAS)"
        },
        { "trait_index": 88, "trait_label": "Erectile dysfunction (ED)" },
        { "trait_index": 71132, "trait_label": "Glycerol levels" },
        { "trait_index": 71131, "trait_label": "IgD on IgD+ B cells" },
        {
          "trait_index": 71126,
          "trait_label": "Immune phenotypes/cell traits (731 traits from GWAS)"
        },
        {
          "trait_index": 71125,
          "trait_label": "Inflammatory cytokines (91 traits from GWAS)"
        },
        {
          "trait_index": 34182,
          "trait_label": "Urokinase-type plasminogen activator (uPA)"
        },
        { "trait_index": 71137, "trait_label": "X-16964 levels" }
      ]
    },
    {
      "pmid": "39744475",
      "title": "Genetically Predicted Immune Cell Traits Mediate the Causal Association Between Plasma Metabolites and Colorectal Cancer.",
      "trait_profile_similarity": 0.6623593948104165,
      "trait_jaccard_similarity": 0,
      "trait_count": 7,
      "involved_traits": [
        { "trait_index": 56281, "trait_label": "16 -hydroxy-DHEA-3-sulfate" },
        { "trait_index": 56283, "trait_label": "CD3 on CD28- CD8+ T cell" },
        { "trait_index": 48, "trait_label": "Colorectal cancer (CRC)" },
        { "trait_index": 9106, "trait_label": "Immune cell traits" },
        { "trait_index": 17152, "trait_label": "Plasma metabolites" },
        { "trait_index": 56282, "trait_label": "SSC-A on CD14+ monocyte" },
        {
```

```
          "trait_index": 56280,
          "trait_label": "Sphingomyelin (d18:1/22:1, d18:2/22:0, d16:1/24:1)"
        }
      ]
    },
    {
      "pmid": "38533503",
      "title": "Unraveling the causal role of immune cells in gastrointestinal tract
cancers: insights from a Mendelian randomization study.",
      "trait_profile_similarity": 0.6614144126122649,
      "trait_jaccard_similarity": 0,
      "trait_count": 16,
      "involved_traits": [
        { "trait_index": 74811, "trait_label": "CCR2 on CD14- CD16+ monocyte" },
        { "trait_index": 74812, "trait_label": "CD19 on IgD+ CD38-" },
        { "trait_index": 74813, "trait_label": "CD19 on IgD+ CD38- naive" },
        {
          "trait_index": 74814,
          "trait_label": "CD25hi CD45RA+ CD4 not Treg AC"
        },
        { "trait_index": 74815, "trait_label": "CD27 on unsw mem" },
        { "trait_index": 74816, "trait_label": "CD28 on CD39+ activated Treg" },
        { "trait_index": 74817, "trait_label": "CD45 on CD4+" },
        { "trait_index": 377, "trait_label": "Colon cancer" },
        { "trait_index": 651, "trait_label": "Esophageal cancer" },
        { "trait_index": 428, "trait_label": "Gastric cancer" },
        {
          "trait_index": 74809,
          "trait_label": "Immune traits - absolute cell (AC)"
        },
        {
          "trait_index": 74807,
          "trait_label": "Immune traits - median fluorescence intensity (MFI)"
        },
        {
          "trait_index": 74810,
          "trait_label": "Immune traits - morphological parameters (MP)"
        },
        {
          "trait_index": 74808,
          "trait_label": "Immune traits - relative cell (RC)"
        },
        { "trait_index": 3298, "trait_label": "Rectal cancer" },
        { "trait_index": 9721, "trait_label": "Small intestine cancer" }
      ]
    },
    {
      "pmid": "39876754",
      "title": "Immune cells: Mediators in the metabolites and Alzheimer's disease.",
      "trait_profile_similarity": 0.659726779569279,
      "trait_jaccard_similarity": 0,
      "trait_count": 14,
      "involved_traits": [
        { "trait_index": 26388, "trait_label": "4-hydroxycoumarin" },
        { "trait_index": 3, "trait_label": "Alzheimer's disease" },
        {
          "trait_index": 48465,
          "trait_label": "CD33dim HLA-DR+ CD11b- myeloid cells"
        },
```

```
        { "trait_index": 48463, "trait_label": "CD64 on monocytes" },
        { "trait_index": 48461, "trait_label": "Glycolithocholate sulfate" },
        {
          "trait_index": 48464,
          "trait_label": "HLA-DR on CD14+ CD16- monocytes"
        },
        { "trait_index": 48466, "trait_label": "HLA-DR on CD8+ T cells" },
        { "trait_index": 48459, "trait_label": "Histidine/pyruvate ratio" },
        { "trait_index": 48460, "trait_label": "Homoarginine" },
        {
          "trait_index": 48462,
          "trait_label": "Immune cell phenotypes (n=731)"
        },
        { "trait_index": 48458, "trait_label": "Metabolites (n=1400)" }
      ]
    }
  ]
}
```

**Evidence profile similarity results (JSON)**

```
{
  "query_pmid": "40325806",
  "query_model": "gpt-5",
  "query_title": "Unveiling the Role of Immune Cells and Plasma Metabolites in Breast
Cancer Risk: A Mendelian Randomization and Mediation Analysis.",
  "query_result_count": 5,
  "similar_studies": [
    {
      "pmid": "38933268",
      "title": "Plasma metabolites as mediators in immune cell-pancreatic cancer risk:
insights from Mendelian randomization.",
      "direction_concordance": 0,
      "matched_pairs": 2,
      "match_type_exact": false,
      "match_type_fuzzy": true,
      "match_type_efo": false,
      "matched_evidence_pairs": [
        {
          "query_exposure": "CCR2 on monocytes",
          "query_outcome": "Breast cancer",
          "query_direction": "decreases",
          "similar_exposure": "CD11c+ monocytes",
          "similar_outcome": "Pancreatic cancer",
          "similar_direction": "increases",
          "match_type": "fuzzy"
        },
        {
          "query_exposure": "IgD- CD38dim AC cells",
          "query_outcome": "Breast cancer",
          "query_direction": "increases",
          "similar_exposure": "HLA DR+ CD8br T cells",
          "similar_outcome": "Pancreatic cancer",
          "similar_direction": "increases",
          "match_type": "fuzzy"
        }
      ]
    },
    {
```

```
      "pmid": "38935111",
      "title": "The association between immune cells and breast cancer: insights from
Mendelian randomization and meta-analysis.",
      "direction_concordance": -1,
      "matched_pairs": 1,
      "match_type_exact": false,
      "match_type_fuzzy": true,
      "match_type_efo": false,
      "matched_evidence_pairs": [
        {
          "query_exposure": "IgD- CD38dim AC cells",
          "query_outcome": "Breast cancer",
          "query_direction": "increases",
          "similar_exposure": "CD3 on CD28+ CD4-CD8- T cells",
          "similar_outcome": "Breast cancer",
          "similar_direction": "decreases",
          "match_type": "fuzzy"
        }
      ]
    }
  ]
}
```


### S7 Case study analyses

#### S7.1 Temporal analysis supplementary materials

Table S7-1: **Temporal evolution of MR research across methodological eras.** Summary of key characteristics across five methodological eras defined by major developments: Early MR (2003-2014), MR-Egger regression (2015-2017), MR-PRESSO outlier detection (2018-2019), within-family MR designs (2020), and STROBE-MR reporting guidelines (2021-2025). Era boundaries roughly align with publication of major methodological papers that shaped the field, though some works might fall outside of the boundaries (e.g. [3]).

| Era | Studies (n) | Year Start | Year End | Mean Traits/Study | Median Traits/Study |
|---|---|---|---|---|---|
| Early MR | 454 | 2003 | 2014 | 4.11 | 4.00 |
| MR-Egger | 631 | 2015 | 2017 | 4.98 | 4.00 |
| MR-PRESSO | 913 | 2018 | 2019 | 5.55 | 5.00 |
| Within Family MR | 821 | 2020 | 2020 | 6.00 | 5.00 |
| STROBE-MR | 12,780 | 2021 | 2025 | 7.14 | 6.00 |

#### S7.2 Reproducibility analysis supplementary materials

Table S7-2: **Concordance distribution statistics by match type and outcome category.** Summary statistics of direction concordance distributions shown in Figure panels C (overall, ”All” rows) and D (category-specific rows). Table presents sample size (N), mean, median, and standard deviation (SD) for each combination of outcome category and match type. Match type definitions: Exact (identical trait descriptions after case-insensitive normalisation, highest confidence), Fuzzy (pairs that failed exact matching but exceeded cosine similarity threshold of 0.70 on 200-dimensional embeddings, with both exposure and outcome required to meet threshold; lower confidence). Statistics correspond directly to ridge plot distributions in Figure 5, panels C and D.

| Outcome Category | Match Type | N | Mean | Median | SD |
|---|---|---|---|---|---|
| All | Fuzzy | 963 | 0.414 | 1.000 | 0.760 |
| | Exact | 261 | 0.739 | 1.000 | 0.441 |
| autoimmune | Fuzzy | 65 | 0.173 | 0.500 | 0.871 |
| | Exact | 41 | 0.843 | 1.000 | 0.309 |
| cancer | Fuzzy | 341 | 0.309 | 0.429 | 0.735 |
| | Exact | 48 | 0.462 | 0.500 | 0.480 |
| cardiovascular | Fuzzy | 299 | 0.494 | 1.000 | 0.730 |
| | Exact | 115 | 0.749 | 1.000 | 0.446 |
| metabolic | Fuzzy | 162 | 0.394 | 1.000 | 0.823 |
| | Exact | 33 | 0.873 | 1.000 | 0.413 |
| other | Fuzzy | 4 | 1.000 | 1.000 | 0.000 |
| | Exact | 1 | 0.500 | 0.500 | NaN |
| psychiatric | Fuzzy | 92 | 0.725 | 1.000 | 0.626 |
| | Exact | 23 | 0.903 | 1.000 | 0.324 |

Table S7-3: **Linear regression model predicting direction concordance.** Ordinary least squares regression coefficients for a model predicting mean direction concordance across trait pairs from publication year, study count, and match type.

| Variable | Coefficient | Std. Error | 95% CI | P-value |
|---|---|---|---|---|
| Intercept | 47.823 | 25.014 | [-1.228, 96.875] | 0.056 |
| Publication year | -0.024 | 0.012 | [-0.048, 0.001] | 0.056 |
| Study count | -0.024 | 0.005 | [-0.034, -0.014] | <0.001 |
| Match type (exact) | 0.327 | 0.025 | [0.278, 0.376] | <0.001 |

*Note:* N = 2,075 trait pairs. $R^2$ = 0.0325, Adjusted $R^2$ = 0.0311. F-statistic = 23.21 (p < 0.001).

Table S7-4: **Ordinal trend test for study count effect on concordance.** Tests for monotonic trend in direction concordance across ordered study count bands (2–3, 4–6, 7–10, 11+ studies). The ordinal trend coefficient represents the change in mean concordance per increase in study count band. Adjusted model controls for match type (exact vs non-exact).

| Test | Coefficient | Std. Error | 95% CI | P-value |
|---|---|---|---|---|
| *Parametric tests* | | | | |
| Ordinal trend (unadjusted) | $-0.056$ | 0.024 | $[-0.104, -0.008]$ | 0.021 |
| Ordinal trend (adjusted) | $-0.092$ | 0.024 | $[-0.140, -0.045]$ | $<0.001$ |
| *Non-parametric tests* | | | | |
| Spearman $\rho$ | $-0.190$ | — | — | $<0.001$ |
| Kruskal-Wallis $H$ | 80.86 | — | — | $<0.001$ |

*Note:* N = 2,075 trait pairs across 4 study count bands. Ordinal encoding: 1 = 2–3 studies, 2 = 4–6 studies, 3 = 7–10 studies, 4 = 11+ studies.

Table S7-5: **Summary statistics of direction concordance by study count band.** Descriptive statistics for mean direction concordance across trait pairs, stratified by the number of studies examining each exposure-outcome relationship. IQR = interquartile range (25th to 75th percentile).

| Study count | n | Mean | SD | SE | Median | IQR | Range |
|---|---|---|---|---|---|---|---|
| 2–3 | 1,492 | 0.50 | 0.80 | 0.02 | 1.00 | [0.00, 1.00] | [−1.00, 1.00] |
| 4–6 | 437 | 0.44 | 0.61 | 0.03 | 0.50 | [0.00, 1.00] | [−1.00, 1.00] |
| 7–10 | 105 | 0.33 | 0.48 | 0.05 | 0.33 | [0.00, 0.71] | [−0.71, 1.00] |
| 11+ | 41 | 0.44 | 0.48 | 0.08 | 0.43 | [0.20, 0.82] | [−1.00, 1.00] |
| Total | 2,075 | | | | | | |

*Note:* Direction concordance ranges from −1 (complete discordance) to 1 (complete concordance). SD = standard deviation; SE = standard error of the mean.

#### S8 Declaration on the use of large language models

In our work, as reported in the main text LLMs are used as part of the research in performing data extraction.

In addition, LLMs have been used as coding agents to assist:

- for research analysis, the implementation of code and documentation;
- for the manuscript, the scaffolding (creation of the LaTeX codebase and manuscript structure), the initial outlining, and the final proof-reading.

Coding agents used in our work include GitHub Copilot (`https://github.com/features/copilot`) and OpenCode (`https://github.com/sst/opencode`). The LLMs used in coding agents include Anthropic Claude Sonnet 4, Anthropic Claude Sonnet 4.5, Anthropic Claude Opus 4.5, OpenAI GPT-4.1, OpenAI GPT-5, and OpenAI GPT-5-mini. The LLMs are accessed via GitHub Copilot and our OpenAI platform account.

Figures 1 and 3 are created by the lead author (Y.L). Other figures and tables used in the manuscript are generated from analysis code (publicly available in Git repositories on GitHub) which AI was used in their implementation, but are not directly generated by AI.

Other non-AI / non-LLM tools in proof-reading the manuscript include textidote (`https://github.com/sylvainhalle/textidote`) and typos (`https://github.com/crate-ci/typos`). The manuscript is primarily written in LaTeX using Neovim (`https://github.com/neovim/neovim`) and we used Overleaf (`https://www.overleaf.com`) for reviewing and co-editing.

The final manuscript is fully drafted by the lead author (Y.L) and has been reviewed and edited by all co-authors. Prior to submission we have reviewed ISCB Policy for Acceptable Use of Large Language Models (`https://www.iscb.org/iscb-policy-statements/iscb-policy-for-acceptable-use-of-large-language-models`) and follow its guidelines on the acceptable uses.

#### References

[1] Yiran Li, Yinfu Zhu, Kunxiang Gong, Yi Wang, Yinger Huang, and Wenbo Hao. Unveiling the role of immune cells and plasma metabolites in breast cancer risk: A mendelian randomization and mediation analysis. *Current Pharmaceutical Biotechnology*, 26, May 2025.

[2] Jianhai Chen, Huabin He, and Ling Gao. Causal role of ischemic heart disease in ovarian cancer subtypes. *Discover Oncology*, 16(1), January 2025.

[3] Neil M Davies, Laura D Howe, Ben Brumpton, Alexandra Havdahl, David M Evans, and George Davey Smith. Within family mendelian randomization studies. *Human Molecular Genetics*, 28(R2):R170–R179, 2019.
